# High-effect gene-coding variants impact cognition, mental well-being, and neighborhood safety substrates in brain morphology

**DOI:** 10.1101/2024.05.21.24307729

**Authors:** Jakub Kopal, Guillaume Huguet, Justin Marotta, Shambhavi Aggarwal, Nicole Osayande, Kuldeep Kumar, Zohra Saci, Martineau Jean-Louis, Xiaoqian J Chai, Tian Ge, B.T. Thomas Yeo, Paul M Thompson, Carrie E Bearden, Ole A. Andreassen, Sébastien Jacquemont, Danilo Bzdok

## Abstract

Our genetic makeup, together with environmental and social influences, shape our brain’s development. Yet, the imaging genetics field has struggled to integrate all these modalities to investigate the interplay between genetic blueprint, environment, human health, daily living skills and outcomes. Hence, we interrogated the Adolescent Brain Cognitive Development (ABCD) cohort to outline the effects of rare high-effect genetic variants on brain architecture and corresponding implications on cognitive, behavioral, psychosocial, and socioeconomic traits. Specifically, we designed a holistic pattern-learning algorithm that quantitatively dissects the impacts of copy number variations (CNVs) on brain structure and 962 behavioral variables spanning 20 categories in 7,657 adolescents. Our results reveal associations between genetic alterations, higher-order brain networks, and specific parameters of the family well-being (increased parental and child stress, anxiety and depression) or neighborhood dynamics (decreased safety); effects extending beyond the impairment of cognitive ability or language capacity, dominantly reported in the CNV literature. Our investigation thus spotlights a far-reaching interplay between genetic variation and subjective life quality in adolescents and their families.

## Introduction

Our genes collectively provide the blueprint of our biological makeup upon which the dynamic interplay of environmental influences, social interactions, and developmental experiences unfolds. The genetic architecture contributes directly and indirectly to the wiring of brain circuits and provides the foundation of behavior repertoire manifestation^1,2^. By understanding genetic underpinnings, we can unravel the origins of individual differences in cognitive processes and behaviors, which in turn offers insights into both adaptive capacities and developmental vulnerabilities^3^. Identifying biological determinants behind brain organization and behavioral differentiation necessitates an integrative approach that cuts across an array of disciplines. Nevertheless. neuroimaging genetics, psychiatric genetics, and environmental factor studies have been conducted in isolated silos. Without the integration of these disciplines, the full extent of genetic influences on human health will remain concealed.

The genetic underpinnings have been traditionally studied through genome-wide association studies (GWAS). However, GWAS have been restricted to common variants which mainly reside in non-coding regions and exert only small effects on phenotypes^4^. Compared to incumbent single nucleotide polymorphism analyses in GWAS, protein-coding copy number variations (CNVs) represent rare and consequential genome-wide perturbations invariably leading to a large decrease or increase in gene expression - a currently under-exploited genetic phenomenon. This class of genetic variation is defined as either a deletion or duplication of sequences of nucleotides more than 1,000 base pairs long^5,6^. Moreover, given recently accumulating evidence, many protein-coding CNVs are now being understood to exert body-wide implications^7,8^ as well as increase the risk of neurodevelopmental disorders, including intellectual disability, autism spectrum disorder or schizophrenia^9–11^. Since protein-coding CNVs are cumulatively relatively frequent in the population and have the potential for substantial effects on a given phenotype, they represent a potent imaging-genetics tool, which we will here deploy for interrogating the effects of genetic modifications on brain physicality and behavioral differentiation in adolescents.

Adolescence is a critical juncture where the seeds of mental health and well-being are sown. During this period, brain circuits and behavioral tendencies undergo dynamic changes shaped by genetic factors, environmental influences, and their interactions^12,13^. Importantly, adolescence is also a life stage during which symptoms of numerous psychiatric disorders become apparent^14^. Recent findings underscore the necessity of adopting a multidimensional and interdisciplinary approach that cuts across sociology, psychology, and biology, conventionally studied in isolation. Such a holistic perspective is essential for a more nuanced understanding of the intricate interplay of genetic, socio-economic, and environmental factors influencing healthy children’s development^15^. By integrating information from cognitive assessments, genetic information, and socio-environmental measures, we can identify potential risk factors as well as unveil protective elements contributing to resilience in individuals navigating the complexities of adolescence^16,17^. Consequently, the analysis of CNVs in adolescents is uniquely positioned to carve out important interactions between our genetic heritage, the environmental milieu, and the intricacies of cognitive and social development, laying the groundwork for a more comprehensive approach to mental health.

In the present study, we leveraged under-studied rare genetic alterations (genome-wide CNVs) with strong downstream effects. We interrogated the Adolescent Brain Cognitive Development (ABCD) study^18^, which represents one of the largest collections of brain images and genetic profiles from over 10,000 children aged 9 to 11 years at baseline. Moreover, these adolescents are prospectively deeply phenotyped by means of an extensive battery of cognitive, behavioral, clinical, psychosocial, and socioeconomic measures. Benefiting from this comprehensive multimodal data, we investigated the effects of a genomic deletion or duplication on patterns of individual subjects’ brain architecture linked with cognitive, behavioral, psychosocial, and socioeconomic measures in a single multivariate analysis. Specifically, we first probed the carefully curated data from 7,657 children for the presence of CNVs. We then deployed multivariate pattern learning algorithms in the children without any CNV to estimate modes of population covariation between brain architecture, represented by 148 regional atlas volumes and ∼1000 behavioral variables spanning 20 rich categories. Finally, we quantified the effects of deletions and duplications on the revealed canonical modes. The robustness of derived modes and CNV-induced differences were assessed by cross-validation and permutation testing^15,19^. This multidimensional and doubly multivariate framework revealing the multi-faceted relationships between genes, brain architecture, and behavior paves the way for innovation in neuroscience, genetics, and personalized medicine.

## Results

### Genome-wide mutations alter patterns of brain and behavior

We used a pattern-learning approach to analyze the impact of genome-wide CNVs in the ABCD cohort by means of its uniquely deep phenotype profiling. To this end, in the group of 7,657 children that passed genetic and MRI quality control, we first identified 514 children carrying at least one genomic deletion and 1,472 children carrying at least one duplication that fully encompassed at least one gene. The 136 children that carried both a deletion and duplication were not analyzed in this study due to the limited sample size. The remaining 73% (5,535) of the children did not carry any protein-coding CNV larger than 50kb across the genome (Fig. 1A). These participant groups (deletions, duplications, controls) showed similar proportions of sex (percentage of females: 44-48%) and distributions of age (Fig. 1A).

**Figure 1.**
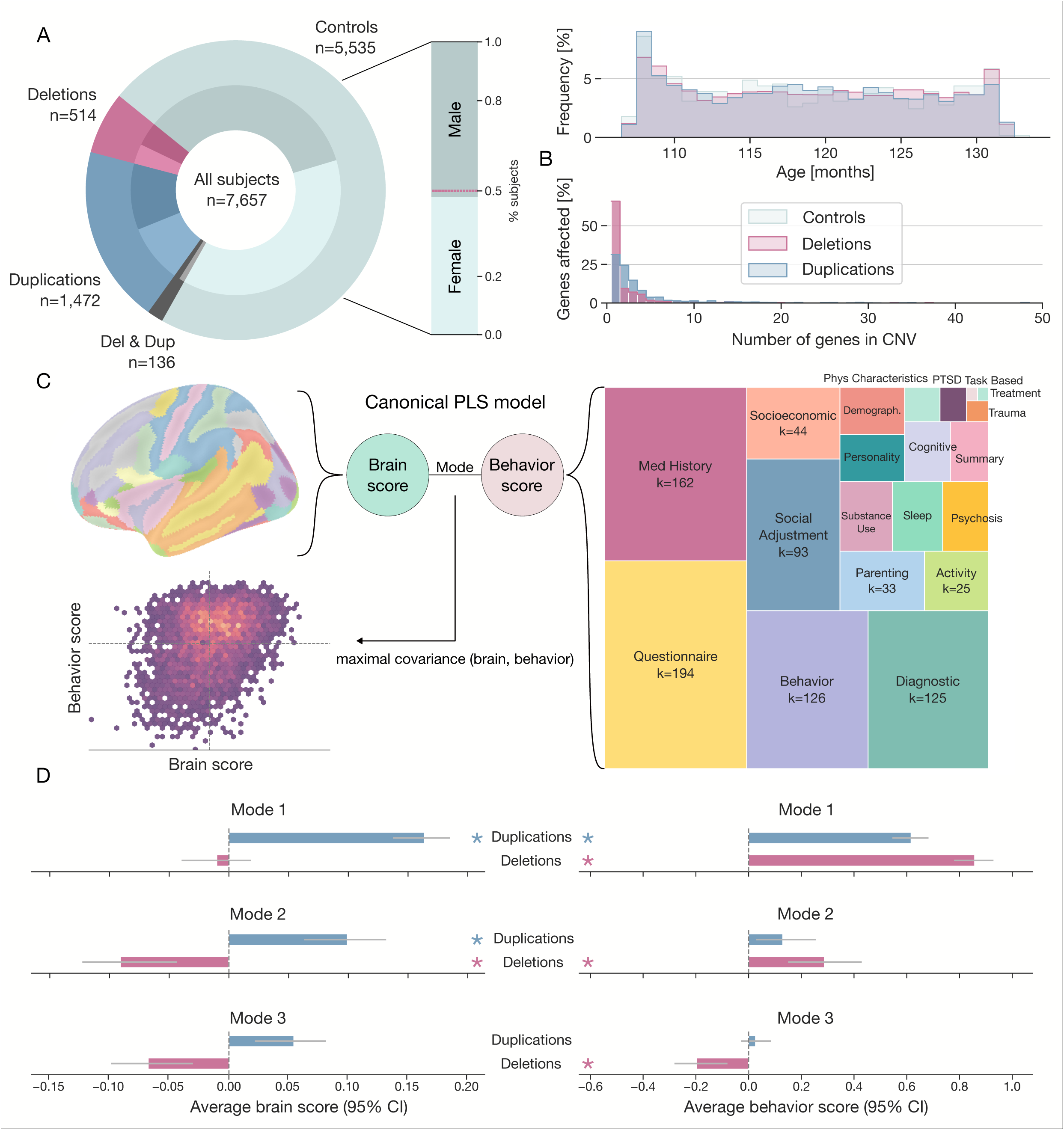
Genome-wide CNVs impact brain-behavior relationships across distinct modes of population covariation. A) Genome-wide CNV identification in the ABCD population cohort. We investigated 7,657 children from the ABCD database. In total, 5,535 children do not carry any protein-coding CNV, 514 carry a deletion and 1,472 carry a duplication fully encompassing one or more genes. 136 participants carried both deletion and duplication. The ratio of males and females is similar in every group (*left plot*, *inner circle*). The age of participants is similar across different CNV groups (*right plot*). B) CNV characteristics. The most common kind of CNV is a deletion/duplication encompassing a few genes. On average, duplications contain more genes than deletions in the ABCD cohort. C) Partial least squares model links the brain with behavior in one holistic model. We estimate a multivariate relationship structure among 148 brain atlas volume measures and ∼1000 behavior measures spanning 20 categories based on measurements from children without any CNV. The canonical scores represent the latent variable expressions calculated from linear combinations of the original brain and linear combinations of behavior measurements that maximize the covariance between the two sets of variables. *k* is the number of phenotypes per category. D) CNV status impacts individual expression strengths of brain and behavior patterns. We compared the average brain and behavior scores for CNV carriers with control participants not used to derive model parameters (i.e., controls not seen by the model during training). Stars denote significant differences based on cross-validation testing (cf. Methods). These results reveal that carrying a CNV significantly impacts canonical scores across different modes of brain-behavior covariation, emphasizing the utility of a multivariate holistic framework that cuts across single disciplines.

Next, we zoomed in on the CNVs that we localized in the children’s genetic profiles (Fig. 1B). Almost 60% of deletions encompassed a single complete gene. Duplications generally encompassed more affected genes than deletions, although a single-gene duplication was the most common (∼30% of cases). Besides the genetic profiling, the ABCD resource provides brain and behavior measurements for each participant: brain measurements were represented by 148 regional brain volumes defined according to the Desikan-Killiany standard atlas. Behavioral measures drew across 962 different phenotypes spanning 20 categories for in-depth follow-up analyses.

In order to investigate how genetic mutations impact brain and behavior, we first established the link between measurements of brain architecture and behavior using a single holistic multivariate model. Specifically, we brought to bear a partial least squares (PLS) model that maximizes the covariation between the weighted set (linear combination) of sociodemographics, family well-being, physical characteristics, or behavioral measures and a weighted set (linear combination) of brain structure measures (Fig. 1C). The PLS model parameters were initially estimated in participants without any CNV, as a reference group, to reveal the modes of covariation that reflect the general population. The subject-wise expressions of each brain-behavior covariation mode (i.e., canonical variables) will henceforth be called *scores*. In other words, these *scores* are calculated as a linear combination (weighted sum) of the original variables with PLS weights. Each identified PLS mode can thus be characterized by a set of brain and behavior scores for all subjects and for each PLS mode. Using a robust protocol for cross-validation and empirical permutation testing^15^, we identified three significant PLS modes (Sup. Fig. 1). The revealed major sources of covariation in adolescents captured the ways in which brain features are intertwined with early life events, mental well-being, or environment.

In the next step, we wished to evaluate if carrying a coding CNV led to statistically significant shifts in the observed brain and behavior patterns. To that end, we devised a bootstrap validation scheme that compares PLS scores between controls and CNV carriers (Sup. Fig. 2). In this CNV-control comparison, deletion and duplication carriers are pitted against control participants not used to derive PLS parameters in order to prevent overfitting (cf. Methods for details). The comparison was based on separately testing the difference in the average behavior scores and the average brain scores. We were thus able to assess mode expression differences separately for each CNV status (Fig. 1C). We found deletions to impact behavior scores in all three identified modes (p-value_mode1_ = 10^-4^, p-value_mode2_ = 0.014, p-value_mode3_ = 0.041). By contrast, duplications affected only the first behavioral mode (p-value_mode1_ = 10^-4^, p-value_mode2_ = 0.151, p-value_mode3_ = 0.336). Furthermore, there was a significant shift in brain scores for the first two modes in duplications (p-value_mode1_ = 10^-4^, p-value_mode2_ = 0.027, p-value_mode3_ = 0.097) and the second mode in deletions (p-value_mode1_ = 0.395, p-value_mode2_ = 0.043, p-value_mode3_ = 0.052). A sensitivity analysis demonstrated that the obtained differences were not driven by the presence of recurrent CNVs, such as 16p11.2 or 22q11.2 (Sup. Fig. 3). These results revealed that carrying a CNV significantly impacts the expression of patterns linking brain architecture and diverse aspects of cognitive, psychosocial, and socioeconomic measures in our ABCD sample. Therefore, genetic factors shape individual differences in brain-behavior correspondences in adolescents.

### Canonical modes bind patterns of whole-brain architecture with real-life functioning, mental well-being and environment

After identifying robust deviations of brain-behavior patterns in CNV carriers at population scale, we examined each revealed mode in more detail. The dominant mode portrayed the ties between large-scale brain networks with sociodemographics and cognition. Specifically, we first re-expressed the difference in PLS scores between controls and CNV carriers using Cohen’s d measure to provide a standardized measure of CNV-carriership effect size. The dominant PLS mode was characteristic of significantly altered behavior scores, with the shift being more prominent in CNV deletions (CNV - controls Cohen’s d_DEL_ = -0.16, d_DUP_ = -0.11) (Fig. 2A). To find which phenotypes play a prominent role in the first mode, we calculated brain and behavior loadings. Our version of these loadings was here obtained by Pearson’s correlation between a respective PLS score and the original measurement. As an example, each brain loading indexes the linear association strength between brain region measurements and brain scores in our reference group. Among the strongest brain effects, we observed the medial orbital sulcus (average of left and right hemisphere Pearson’s r = 0.25), a part of the frontal lobe which may be involved in various cognitive functions, including decision-making, emotional processing, and social cognition ^20^. Since duplication carriers displayed higher brain scores compared to controls and since medial orbital sulcus was associated with positive loading (higher volume = higher score), this result pointed to increased volume in this region for duplication carriers. Other strong negative loadings included the middle occipital sulcus, subcallosal area, superior occipital gyrus, or right lingual gyrus (Fig. 2B). We subsequently submitted these loadings to a bootstrap test to determine if they were significantly different from zero (cf. Methods). This test was based on 1,000 PLS model instances built on a randomly perturbed version of our ABCD participants created by sampling a participant cohort of the same sample size (with replacement). We further computed the relative number of significant region effects (i.e., loadings) among each set of regions belonging to one of the seven large-scale networks according to Schaefer-Yeo definitions. We observed that at least 50% of the loadings in each of the seven networks were significant, highlighting the brain-wide pattern of this first mode.

**Figure 2.**
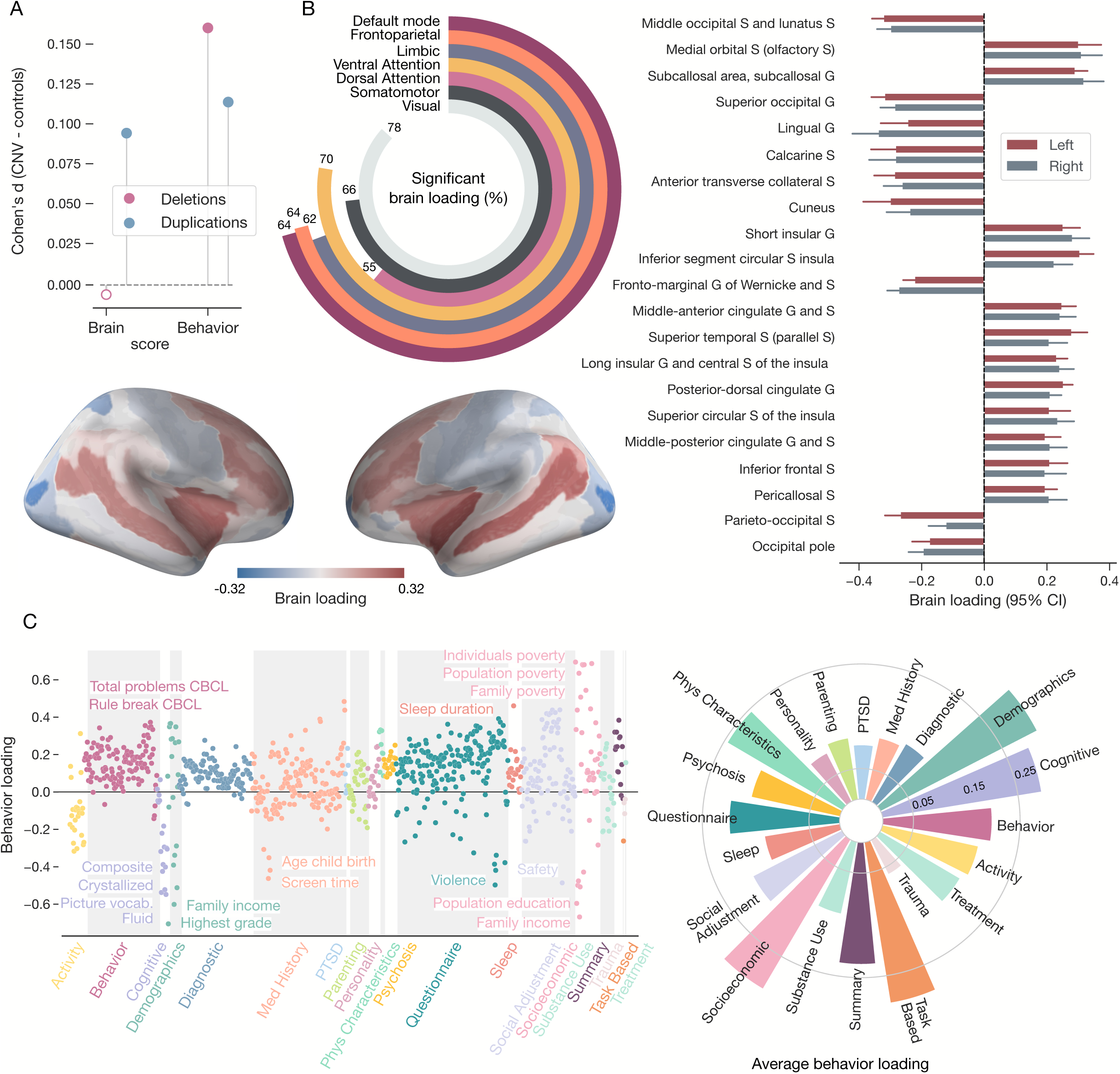
The leading population mode tracks decreased cognitive functioning in CNV carriers. A) CNVs significantly impact revealed dominant behavior pattern. Cohen’s d values of canonical scores calculated between controls and separately carriers of deletions and duplications are plotted for the first canonical mode. Both deletion and duplication carriers show a significant shift in behavior scores, with a stronger effect for deletions. Only duplications display significantly affected brain scores. Significant differences are represented by solid dots. B) Brain region correlates reveal a whole-brain pattern. Brain loadings are calculated as the correlation between brain scores and 148 regional brain volumes. The bar plot depicts 20 regions with the strongest brain loadings, including a 95% confidence interval based on the bootstrap resampling. G denotes gyrus, and S denotes sulcus. The radial bar chart shows the percentage of significant brain loadings in each of the seven large-scale networks based on the bootstrap significance test. C) Behavior correlates highlight real-life functioning. Behavior loadings are calculated as the correlation between behavior scores and ∼1000 behavior measures. The strongest behavior loadings include a variety of cognitive scores (i.e., summaries of tasks focused on language and vocabulary comprehension, working memory, abstract reasoning, or problem-solving), parental education, or family income. On average, the demographic, socioeconomic, and cognitive categories display the strongest coefficients. In summary, the first canonical mode highlights the connection between most of the brain networks and assessments of cognition and demographics.

Furthermore, we inspected a broad portfolio of behavior characteristics interlocked with the above-described brain-level effects. To that end, we calculated behavior loadings similarly to brain loadings. The strongest loadings included family income (Pearson’s r = 0.70), poverty index (Pearson’s r = -0.68), parental education (Pearson’s r = 0.60), measures of cognitive performance (Pearson’s r = - 0.57), and also screen time or sleep duration (Pearson’s r = 0.46) (Fig. 2C). To obtain a synopsis of the dominant behavioral profile, we averaged absolute behavior loadings in each of the 20 categories. Demographics, cognitive, and socioeconomic categories had the strongest average loadings (average absolute Pearson’s r > 0.22). Since CNV carriers displayed higher expression compared to controls for this mode characterized by negative loading for measures of cognitive performance (lower performance = higher score), these results thus point to decreased cognitive abilities and real-life functioning, especially in deletion carriers. Collectively, the dominant canonical mode highlighted the crosslinks between most of the brain networks and assessments of cognition and demographics.

The second PLS mode spotlighted opposing gene dosage effects on the brain structure that we identified to tie into family history of mental health. Specifically, we observed significant opposing brain average expressions for both deletions and duplication (Fig. 3A), which might reflect the mirroring effect on brain architecture previously reported for CNVs at specific genomic loci^21^. Only behavior scores in deletion carriers passed the significance testing. According to our obtained brain loadings, the mirroring brain scores were mainly driven by the lingual gyrus (across-hemisphere average Pearson’s r = -0.38), followed by supramarginal, precentral, or lateral superior temporal gyri (Fig. 3B). Despite being part of distinct brain networks, these regions were previously associated with neural mechanisms supporting complex cognitive tasks, especially those involving semantic processing or executive functions^22,23^. Collectively, following the conducted bootstrap significance test, most of the significant regions belonged to dorsal attention, somatomotor, and frontoparietal networks (more than 67% significant regions in each network). The interactions and coordinated activity of these networks are known to be essential for the efficient integration and execution of complex cognitive and motor tasks^24^.

**Figure 3.**
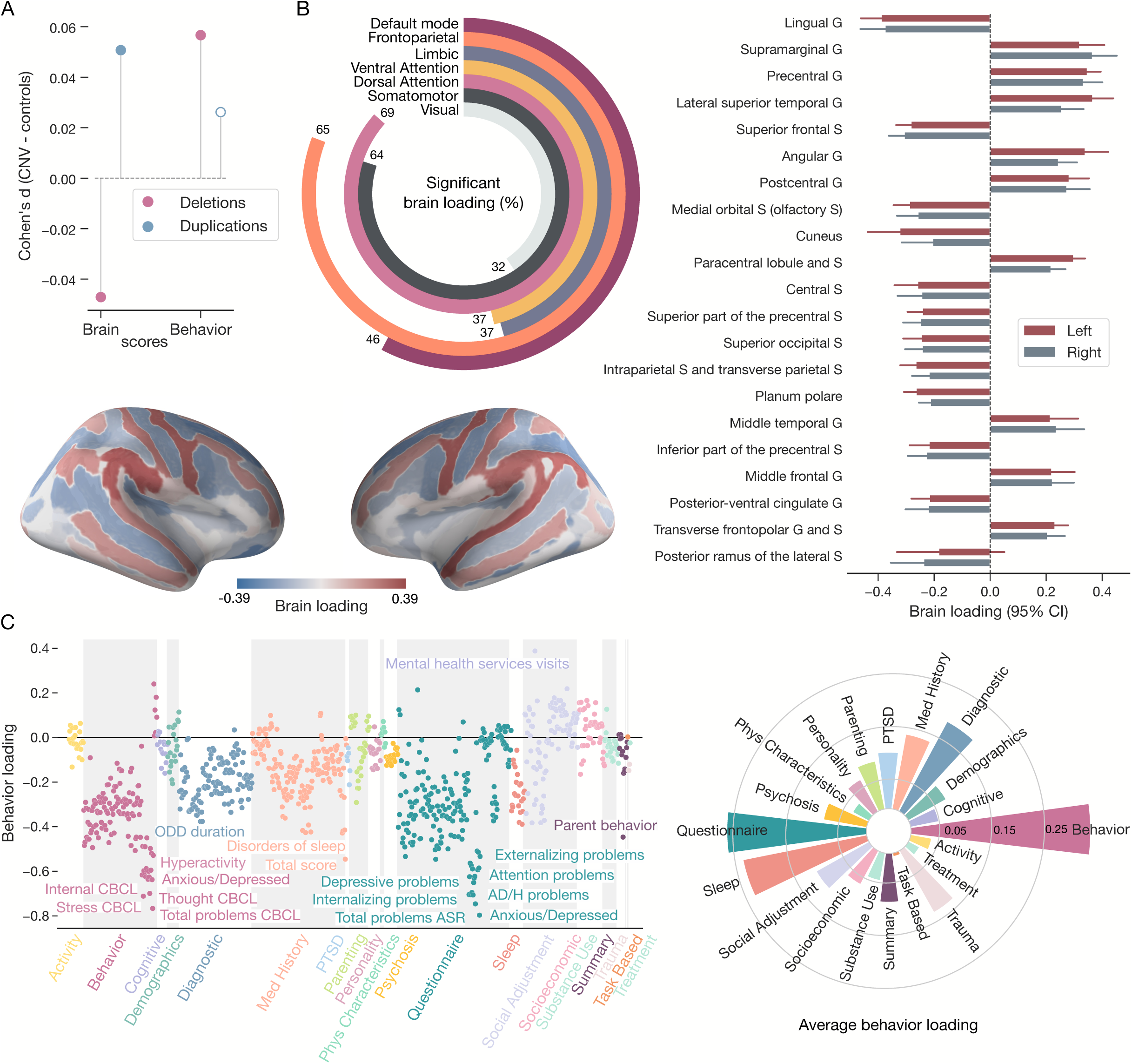
The second population mode spotlights shift in brain scores associated with mental wellbeing. A) Canonical scores untangle gene dosage-induced differences and similarities. Deletions and duplications lead to mirrored significant brain scores. On the other hand, only deletions display significantly affected behavior scores. Significant differences are represented by solid points. B) Three large-scale brain networks dominate brain loadings. The lingual gyrus (G=gyrus, S=sulcus) displays the strongest loading of all regions. Summarized by large-scale networks, the highest percentage of significant brain loadings is present for the dorsal attention, somatomotor, and frontoparietal networks. C) Behavior loadings stress the importance of mental well-being. The strongest behavior loadings include parental and child-reported levels of problems, stress, anxiety, and depression. Therefore, this mode was dominated by mental well-being phenotypes mainly from parental behavior, child questionnaires, and sleep categories based on the average absolute loadings. Collectively, the second canonical mode proposes decreased mental well-being as a significant marker of deletion carriers.

The prominent deviations in behavior scores in deletion carriers can be explained by elevated assessments of mental well-being as revealed by behavior loadings. Specifically, phenotypes from the Child Behavior Checklist (CBCL) and the Adult Self Report (ASR) dominated the set of relevant behavior loadings (Fig. 3C). Particularly, the total scores of CBCL (Pearson’s r = 0.80) and ASR (Pearson’s r = 0.77) emerged as the two strongest loadings. They were followed by measures of both parental and child anxiety, stress, depression as well as child sleep disorders. Indeed, when averaged across categories, the sleep category joined child behavior and parental questionnaires as the most prominent (average Pearson’s r = 0.22). The combination of flagged phenotypes from both children and adult assessments suggests that the second mode captures a comprehensive view of the well-being intricately tied to the family system. In addition, it points towards potential dynastic effects, i.e., the impacts of (inherited) genetic variants on family environments. Collectively, the second canonical mode proposed decreased familial mental well-being as a prominent marker of deletion carriers.

In the third and last canonical mode, we observed the relationship of the default mode and frontoparietal networks with environmental measures. Despite the mirrored effects on brain structure, the only significant shift was found for behavior scores in deletion carriers (Fig. 4A). The third mode was characterized by a strong contribution of the insula (average Pearson’s r = 0.33) as well as middle temporal and lateral superior temporal gyri (Fig. 4B). The majority of significant regions based on bootstrap testing was part of the default mode network or the frontoparietal network (23% and 20% of regions, respectively). These two networks belong to the multimodal end of the unimodal-to-multimodal characterization of large-scale brain networks. Prior research suggests that the relevance of these two networks and underlying regions could imply their crucial roles in cognitive flexibility and the integration of thought processes necessary for problem-solving and decision-making^25^.

**Figure 4.**
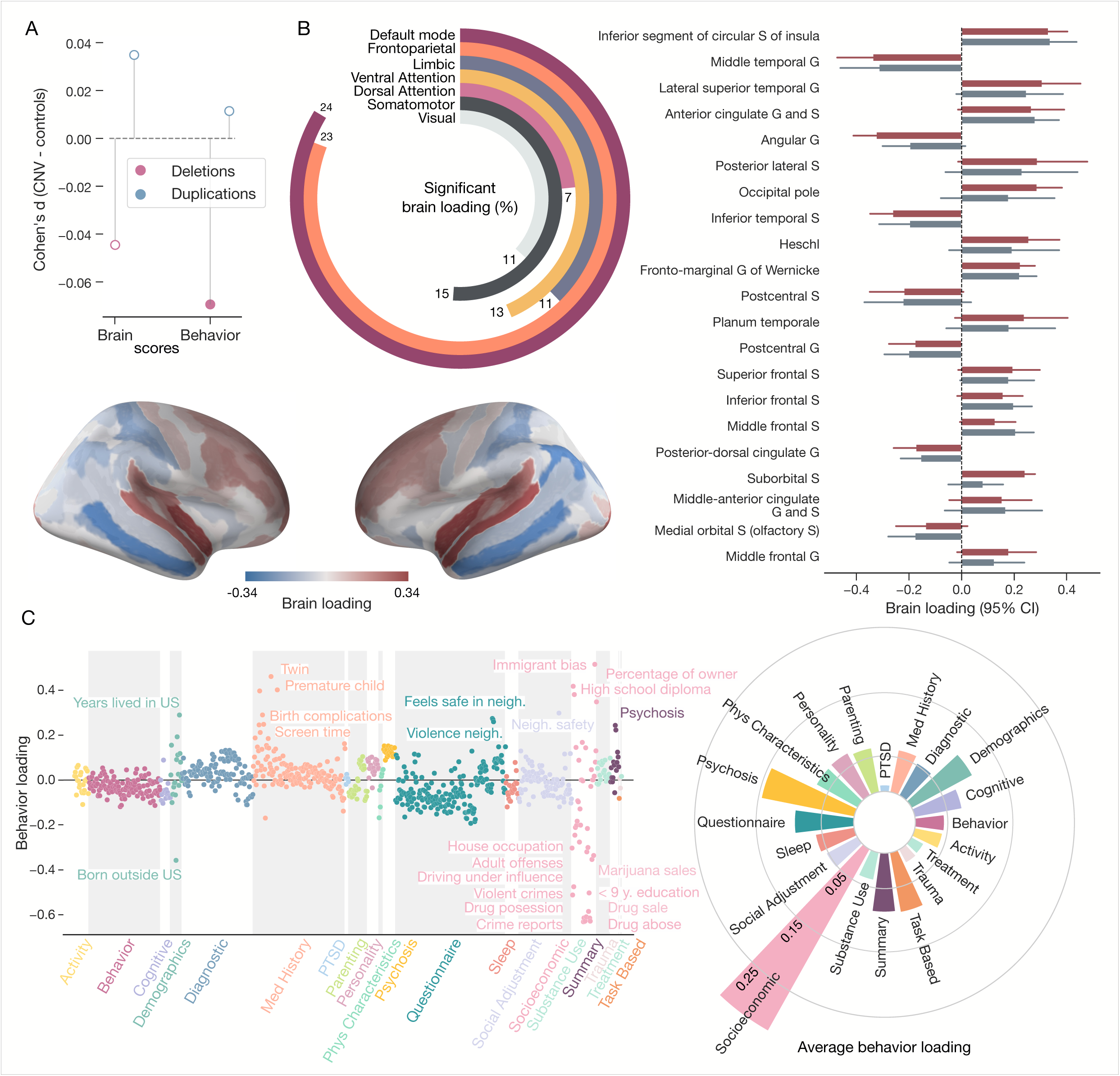
The third population mode links higher order networks to environment measures. A) Deletions, unlike duplications, shape behavior scores. In the third canonical mode, the only significant effect is for behavior scores in deletion carriers. Significant differences are represented by solid points. B) Higher-order networks play a prominent role in the third canonical mode. The anterior insula, followed by the middle temporal gyrus (G=gyrus, S=sulcus), are regions most strongly associated with brain scores. Furthermore, the default mode network, along with the frontoparietal network, displays the highest percentage of significant regions based on the bootstrap significant test. C) Environmental variables characterize behavior loadings. Measures of neighborhood violence, safety and crime are among the strongest loadings. These environment-associated phenotypes come primarily from the socioeconomic category. The third significant mode illustrates how deletions shift the expression of the mode linking the environment and higher-order networks.

Examining the behavior profile in the third canonical mode highlighted variables associated with the environment (Fig. 4C). Concretely, phenotypes related to the neighborhood, such as crime reports (Pearson’s r = 0.63), drug possession, violent crimes, adult offense, and feelings of safety emerged as strongly associated. These phenotypes reflect social and community dynamics, which might affect the overall quality of life for individuals within that context. In summary, the third significant mode revealed an alteration in how environmental differences link to higher-order networks in adolescent deletion carriers.

### Decoding brain and behavioral patterns: Population, genetic and temporal perspectives

After describing how genetic mutations influence the expression of behavior patterns, we explored how characteristics of genes encompassed in CNVs shaped behavior scores. In other words, for each individual with a deletion or duplication, we scored the genes inside a CNV using a total of seven complementary descriptions, including the average temporal expression, number of genes preferentially expressed in the brain, number of genes associated with autism spectrum disorder, schizophrenia, or broader portfolio of disorders, and a functional intolerance score: loss-of-function observed/expected upper bound fraction (LOEUF). LOEUF score provides insights into whether “Loss of function” variants in a given gene are under negative selection pressure in a population. It reflects the gene’s functional impact by indicating how likely the gene will lead to disruptive changes when it is inactivated by a genetic variant. We then performed an exploratory analysis using Pearson’s correlation between behavior scores and the quantitative descriptions of CNVs occurring in the genome (Fig. 5A). For deletions, the strongest observed association was with gene brain expression (Pearson’s r = -0.10, p_FDR_ = 0.13). For duplications, the strongest observed association was with temporal gene expression (Pearson’s r = -0.11, p_FDR_ = 0. 0006). This result suggested deteriorating impact of altering dosage in genes expressed early during the human development. Other strongly associated description was the sum of LOEUF but, due to the limited number of CNV carriers, only the association with genetic temporal profile reached significance after applying False Discovery Rate (FDR) correction to control for multiple comparisons. Nevertheless, the reported associations can serve as valuable pointers for further research. In summary, our findings underscore the intricate relationship between genetic characteristics and behavioral outcomes, highlighting the importance of considering both genetic and temporal dimensions in understanding the etiology of behavioral patterns and susceptibility to disorders.

**Figure 5.**
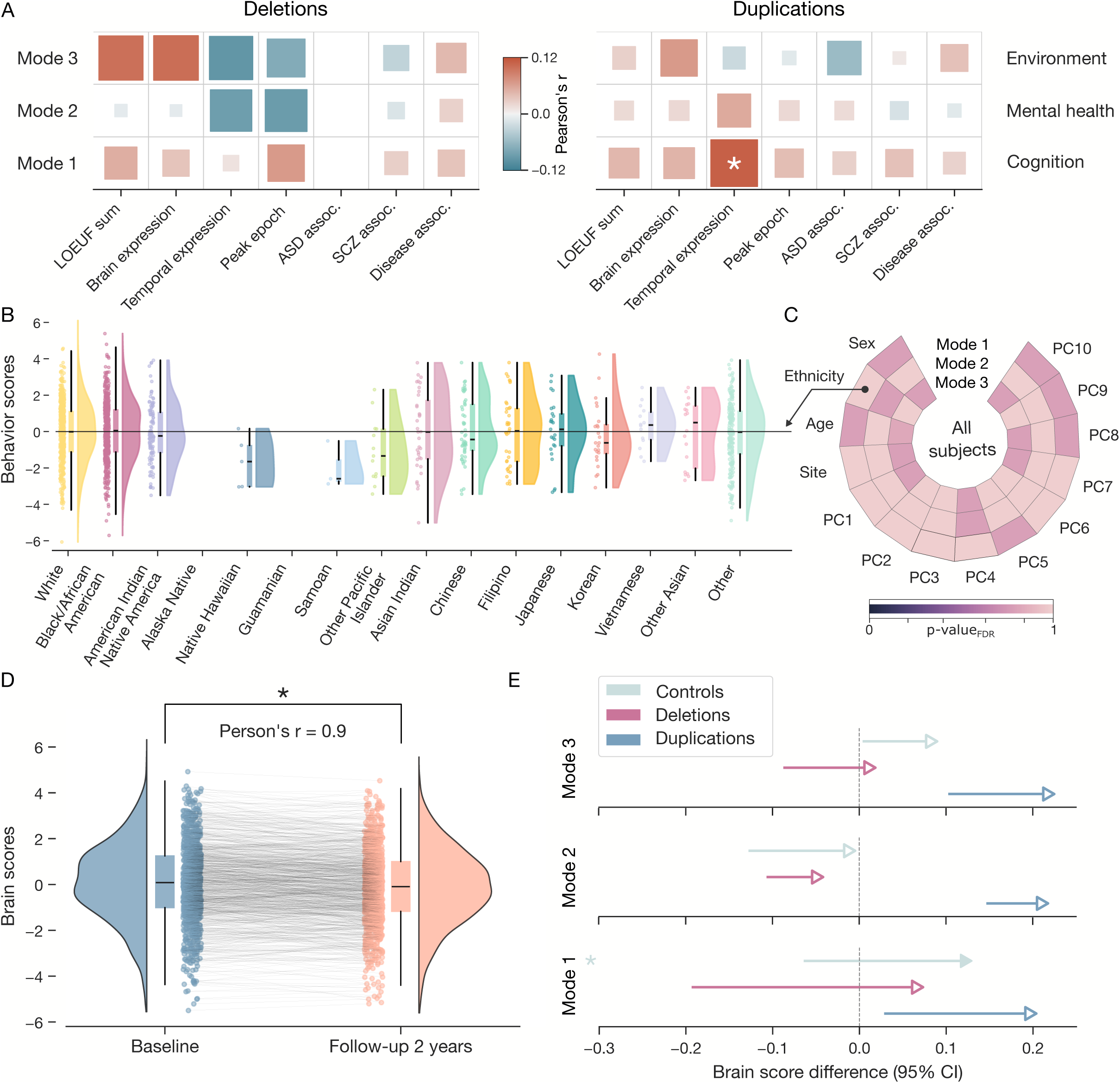
Population modes are driven by temporal gene characteristics rather than sociodemographic factors. A) Behavior scores driven by temporal and spatial expression. Genes encompassed in each CNV were characterized based on seven different metrics. Summaries across all the deleted/duplicated genes for each CNV carrier were used to provide subject-specific annotations. Across the three different modes, behavior scores are most strongly associated with temporal expression, brain expression, and LOEUF scores. The three modes are labeled based on their dominant phenotype. * denotes significant association after FDR correction. B) Behavior scores not explained by ethnicity. As an example, behavior scores of the first canonical model are plotted for all subjects separated by participant ethnicity. The rain cloud plot combines a scatter plot, a box plot (whiskers equal to 1.5 times the interquartile range), and a violin plot. There is no significant difference in mean score expression across the 16 ethnicities. C) Canonical modes not reflective of sociodemographic factors. In addition to ethnicity, behavior scores across the three analyzed modes do not significantly differ based on sex, age, site, and the first ten genetic components capturing major patterns of genetic variation. D) Canonical modes capture brain maturation. A single PLS model evaluated brain measurement from the baseline and 2-year follow-up measurements. The star denotes significant change based on a bootstrap mean test. Both measurements are highly correlated. E) Similar cortical aging across all 3 groups. As an extension, we examine the difference between baseline and 2-year follow-up measurements for all CNV groups and all three canonical modes. The arrow direction symbolizes the direction of change. Significant changes are represented by solid markers and a star. Both CNV groups display similar patterns of brain aging compared to controls, with particular change significant for the first canonical mode.

We then explored whether modes of population stratification, that is, specific sociodemographically defined groups, also influence the derived patterns. In other words, we quantified if ethnicity, sex, age, or genetic background are linked with the shifts in behavior scores. As a concrete example of this sensitivity analysis, we stratified participant-wise behavior scores for the first mode by reported ethnicity (Fig. 5B). Using one-way ANOVA, we assessed whether there were significant differences in scores as a function of these diverse ethnic categories. Notably, the findings revealed that the scores did not exhibit a statistically significant difference among ethnicities (F-statistic = 0.89, p-value = 0.56). We then extended this post-hoc analysis to other modes of population covariation and other metrics of population stratification. Namely, we quantified the difference in scores between males and females using a two-sample t-test and as a function of the 21 recruitment sites using one-way ANOVA. Moreover, we probed the association of scores with age and the ancestry structure of the cohort measured using the first ten principal components of genotyping data. We collected all p-values and applied FDR correction to control for multiple comparisons across the totality of 52 performed tests. None of the performed tests revealed significant association (Fig. 5C). This comprehensive examination provided valuable insights into the potential universality of the observed scores among modes of population stratification, underscoring the importance of considering the generalizability aspect in the broader context of the study’s implications^26^.

As the final step, we extended our analyses by examining longitudinal changes in brain structure between controls and CNV carriers at the two-years-after imaging time point. Investigating the trajectory of brain development over time can provide insights into whether individuals with CNVs exhibit distinct patterns of structural change. We benefited from the availability of 3,654 brain scans measured two years after the first visit (48% of participants passing genetic quality control). We observed a high correlation between regional volumes acquired at these two time points (Sup. Fig. 3). We used a single PLS model that subsequently evaluated brain measurement from the baseline and 2-year follow-up (cf. Methods). Put differently, a single PLS model provided a holistic summary of brain maturation by calculating a brain score for each participant in each visit across the three modes. As an example, similar to brain structure measurements, we observed a strong link between PLS scores in the dominant mode between the baseline and follow-up measurements in controls (Pearson’s r = 0.90) (Fig. 5D). Furthermore, we used a bootstrap mean test (cf. Methods) to evaluate temporal change in brain scores in controls and CNV carriers across the three canonical modes (Fig. 5E). We found a significant decrease in the mean brain score only for the first canonical mode in controls (p-value_FDR_ = 0.001). Nevertheless, both CNV groups displayed similar brain maturation patterns compared to controls. Given the observed similarity in brain structure developmental patterns of CNV carriers and controls, further exploration of earlier stages of life may provide valuable insights into distinctions in neurodevelopmental processes.

## Discussion

In the current study, we carefully examined the ramifications of carrying an exonic CNV on brain organization and behavior. To this end, we designed an analytic framework based on a single holistic pattern-learning algorithm that can quantitatively dissect the impact of genetic mutations on a multimodal measurement that cut across disciplines to untangle the complex genes-brain-behavior interplay. This multivariate model uncovered three significant modes of brain-behavior covariation. The first mode connected volumetric differences in more than 50% of brain regions with measures of cognition and demographics. The second mode linked dorsal attention, somatomotor, and frontoparietal networks with mental health measures. Finally, the third mode highlighted associations between the higher-order networks and environmental factors. We then drew a detailed picture of how carrying a genomic deletion or duplication influences the expression of these comprehensive brain and behavior patterns. Specifically, both classes of CNVs were linked to adverse impacts on family well-being, as seen in the adverse effects on cognitive functioning, mental health, and socioeconomic outcomes. Our results also highlight the similar ramifications for cognition and behavior associated with deletions and duplication despite their distinct effects on brain anatomy, corroborating some of our earlier CNV-imaging studies on the UK Biobank^8,27^. Finally, we demonstrated that different genomic characteristics drive these shifts in behavior differentiation and brain maturation.

Our results call for broadening the scope of genetic analyses in human health beyond what is traditionally considered. However, the analyses of genetic influences have long been dominated by univariate frameworks^28,29^. These standard regression approaches model one outcome variable at a time and thus focus on individual variables independently while neglecting the complex relationships and synergies that exist among genes, brain, and behavior. In other words, univariate approaches struggle with high-dimensional data and are prone to the "curse of dimensionality", making it challenging to capture the joint influence of multiple variables^30^. Scientists have long been waiting for multimodal datasets with deep phenotypic profiling of each participant. However, the availability of such resources prompts a change in our analytic toolkit^31^. PLS addresses several limitations of mass univariate approaches by embracing a doubly-multivariate strategy, providing a more nuanced and integrated perspective on the relationships between thousands of measures of brain architecture and behavior in the general population^15,19^. Prior research showed that genes are an important contributor to the interindividual variability of thus uncovered latent patterns^32^. Building on the heritability of the latent patterns, we showcased that their expression is further shaped by the presence of genome-wide protein-coding mutations, demonstrating their effects on social, familial, and environmental factors.

The consequences for various aspects of human health and well-being often go unnoticed because analyses of genomic deletions and duplications most commonly focus on intellectual disability and developmental delay^33,34^. Developmental delay phenotypes, especially language and motor disorders, are the earliest symptoms for which children are clinically referred for assessments and genetic testing^35^. Recent results showcased potential lifelong implications represented by diminished academic qualifications, occupation or household income for a small set of schizophrenia-associated CNVs^36^. As an extension, our results demonstrated that the genome-wide presence of any coding CNV could lead to impaired real-life functioning, represented here by cognitive performance, income, education, screen time, or sleep duration. These characteristics played a driving role in our dominant mode of population covariation. A similar dominant mode characterized by cognitive measures as well as screen time was identified in the HCP population resources^19^. The stronger influence of deletions on the dominant mode compared to duplications is concordant with the more pronounced effect of deletions on cognitive ability observed in clinical studies^37^. Therefore, the validity of the PLS-derived dimensions is corroborated by recapitulating key findings from previous clinical studies.

The power of our doubly-multivariate approach allowed us to reveal additional consequences beyond just the dominant population mode, which are at risk of staying hidden in classical analyses^38^. Concretely, our second mode highlighted impoverished familial mental well-being as a prominent marker of deletion carriers. Notably, the presence of phenotypes from both child and parental questionnaires demonstrates how well-being is closely tied to the family system as a whole. It has been estimated that over 99% of CNVs are inherited, whereas others arise de novo^39^. Therefore, in addition to influencing offspring phenotype through genetic inheritance, the parental genotype can indirectly influence offspring phenotype through its expression in the parental phenotype^40^. In other words, carriers of the gene-dosage alterations may show certain characteristics similar to the parent’s condition. Where this occurs, offspring may be subject to both phenotype-associated CNV and phenotype-associated environments from parents. These dynastic effects, i.e. “inheritance” of the environment in addition to genotype, might explain how CNVs associated with difficult behavior in the parent’s generation create unfavorable environments, which might inflate behavioral problems in children. In conclusion, the CNVs we studied in adolescents are likely to have been passed down from either parent, which points toward influences on the overall family system. The multigenerational impact where genetic and also environmental legacies contribute to the behavioral outcomes highlights the complex interplay between inherited genetic variations and the environments shaped by parental phenotypes.

Finally, our analyses also spotlighted genomic deletions as shifting the expression of the mode linking the environment and higher-order brain networks. Interestingly, the higher association cortex, especially the default mode network, was suggested to be more “life wired” – resulting from differences in the circumstances and contexts in which people grow up and everyday life experiences^41^. In other words, the deeper layers of the neural processing hierarchy, such as the default mode network, allow for greater environmental influence and plasticity, as demonstrated by prolonged maturation and slower myelination compared to sensory/motor circuits in human primates^42,43^. Our finding adds more evidence for the adaptive and dynamic nature of the recently evolved parts of the human brain, emphasizing the significant role of genetic and environmental interplay in shaping neural development and function. Importantly, the environmental milieu, here represented by measures of crimes, drug possession, or violence, is related to health through psychological, physiological, and behavioral pathways^44^. Prior research documented chronic health conditions to be more prevalent in low-income neighborhoods, including those affecting infants (low birth weight), children (asthma), and adults (cardiovascular health)^45^. Specifically, low socioeconomic status neighborhoods^46^ and neighborhoods perceived as unsafe^47^ displayed elevated physiological risk, which includes indicators of inflammation and neuroendocrine and cardiovascular functioning. Our study thus implies that genetic underpinnings that govern neurodevelopmental trajectories may potentially magnify the impact of environmental stressors on long-term health disparities.

The effects of protein-coding alterations are not limited to a single brain circuit, such as the default mode network. Instead, we painted a detailed picture of effects on brain patterns spanning the entire cortex. In concordance, a previous study identified brain-wide patterns of regional alterations that robustly differentiate controls from carriers of clinical CNVs^8^. Here, we broaden the incumbent analysis scope of a few selected CNVs towards any coding CNV present in the genome. We observed opposite effects of deletions and duplications, recapitulating the mirroring effects observed in clinical studies^21^. As a primary example, the lingual gyrus played a dominant role in two altered brain patterns. The effects of CNVs on this region have been documented for carriers of 16p11.2 CNVs^48^. Similarly, as prominent examples from the frontal lobe, we observed significant contributions of the middle and superior frontal gyrus, which have been shown to be impacted by 1q21.1^49^, resp. 15q11.2^50^ alteration. Impairments of lingual and frontal gyri have been associated with anxiety-depression severity^51^ or attention control deficit^52^ – phenotypes often present in CNV carriers^53^. Though these examples are region-specific, our research indicates far-reaching implications across entire networks. While the identified whole-brain patterns represent a general trend in each class of genetic mutation, the specific alterations pertinent to a specific CNV (e.g., 22q11.2 or 16p11.2) are further molded by the attributes of genes that are affected by a given CNV. According to analyses, the final brain and behavior profile is shaped by various attributes, including the number of deleted/duplicated genes, their tolerance to being mutated, or the temporal expression profile of affected genes. This may be part of the reason why prior research found brain patterns associated with deletions at 22q11.2 loci to strongly resemble deletions at 15q11.2 loci while being different from 16p11.2 deletions^8^. In other words, the here-revealed brain modes represent the bedrock of the global CNV effect upon which the specific CNV effects unfold guided by the properties of encompassed genes, such as temporal and spatial expression profiles.

In conclusion, we developed a multi-level pattern-learning framework to investigate the effects of genome-wide protein-coding mutations on brain organization and behavior. This approach offers a comprehensive view of the multifaceted impact of rare genetic variations, surpassing important limitations of traditional univariate frameworks. We revealed that both genomic deletions and duplications can lead to a toll on family well-being through associations with increased parental and child stress, anxiety and depression, as well as neighborhood violence. This demonstrates that the consequences of genetic mutations are far more extensive than just cognitive functioning, playing a significant role in shaping socioeconomic and environmental aspects of life experiences. Future research building on our multidisciplinary approach can better appreciate the complexity of the relationship between genetic determinants and human health.

## Methods

### ABCD population data source

Brain imaging, behavioral, clinical, and genetic data in this study were obtained from the Adolescent Brain Cognitive Development Study (ABCD; https://abcdstudy.org/), representing the most extensive biomedical child development study of its kind. The ABCD Study acquired data from 11,877 children aged 9–10 years (mean age = 9.49 years) from 21 sites across the United States (48% girls; 57% Caucasian, 15% African American, 20% Hispanic, 8% other)^54^. We leveraged baseline measurements from ABCD Annual curated release 4.0 (https://data-archive.nimh.nih.gov/abcd). Data Release 4.0 contains baseline data on the entire participant cohort as well as early longitudinal data, including 2-year follow-up neuroimaging data (second brain imaging timepoint). All protocols for ABCD are approved by either a central or site-specific institutional review board committee^55^. Caregivers have provided written, informed consent and children provide verbal assent to all research protocols^56^. Additional information about the ABCD Study can be found in^57^. This dataset is administered by the National Institutes of Mental Health Data Archive and is freely available to all qualified researchers upon submission of an access request. All relevant instructions to obtain the data can be found at https://nda.nih.gov/abcd/request-access.

### Genetic annotation and CNV calling

Our study is built on the identification of exonic CNVs in the ABCD study sample. The genotyping protocol for the ABCD sample has been described previously^58^. In addition to the QC provided by ABCD, we performed several additional steps to ensure high quality of the genetic data. Using PLINK v1.9^59^, we removed SNP variants with a missing rate > 5% as well as SNPs with a Hardy-Weinberg equilibrium exact test p-value < 0.0001. We only considered arrays with call rate ≥ 99 %, log R ratio SD < 0.35, B allele frequency SD < 0.08, the absolute value of wave factor < 0.05 and the count of all unfiltered CNV per sample ≤ 10. We also removed subjects with > 5 % missing rate (n = 73) and, all individuals with duplicated data (n = 419, high degree of identity-by-descent, PI_HAT > 0.8) or with discordant phenotypic and genetic information regarding sex (n = 0).

The identification of CNVs using SNP array (GRCh37/hg19) data followed previously published methods^34,60^. CNVs were called using the pipeline described at https://github.com/labjacquemont/MIND-GENESPARALLELCNV. In short, we computed (PFB)-files (Human Genome Build NCBI37/hg19) based on 500 best arrays in ABCD, and we used GC (content)- model files (https://kentinformatics.com and https://github.com/ucscGenomeBrowser/kent.git). Only autosomal CNVs detected by both PennCNV^61^ and QuantiSNP^62^ were used, to minimize the number of potential false positives. All identified CNVs met stringent quality control criteria: confidence score ≥ 30 (for at least one of the two detection algorithms), size ≥ 50 kb, unambiguous type (deletion or duplication), overlap with segmental duplicates, and HLA regions or centromeric regions < 50 %. Finally, all carriers (n = 1) of a structural variant ≥ 10Mb, a mosaic CNV or a chromosome anomaly (aneuploidy or sexual chromosome anomaly) were removed. For the final set of participants, we calculated the first 10 genetic principal components (PCs) using the --pca function from PLINK v2.3^63^.

All identified CNVs were annotated using Gencode V19 (hg19) with ENSEMBL (https://grch37.ensembl.org/index.html). In this study, we only used exonic CNVs that fully encompassed at least one gene. In addition to the number of encompassed genes, each CNV was further annotated with seven other previously used scores. Specifically, we used an annotation quantifying the tolerance to protein-loss-of-function of each gene: Loss-of-function Observed/Expected Upper bound Fraction -LOEUF^64^. Each CNV was then characterized by the sum of LOEUF of encompassed genes. Furthermore, CNVs were described using average temporal expression^65^ and average peak epoch. Gene-wise temporal expression was calculated as the developmental trajectory that the gene follows based on trajectory analysis (gene-specific trajectory coding: ‘Rising’ = 1, ‘Non-transitional’ = 0, ‘Falling’ = -1). The peak epoch corresponds to an epoch of highest expression, where epochs correspond to the developmental period defined previously^66^. Each CNV was also characterized by the number of genes, which expressions in the brain were labeled as ‘High’ or ‘Elevated’ according to the GTEx resource (https://www.gtexportal.org). Finally, we quantified how many genes in each CNV were previously associated with autism spectrum disorder (risk genes from ref. ^67^, schizophrenia (risk genes from ref.^68^, and any disorder by either rare or common variation^65^. The similarity of the seven annotations is summarized in Sup. Fig. 5.

As part of our sensitivity analyses, we compiled a list of 85 CNVs previously proposed to be pathogenic^10,33,69–71^ (sum of 1/LOEUF for each gene encompassed in CNV ≥ 6 or inclusion in ClinGen resource^72^). Regional coordinates are available in ref. ^34^. CNV was defined as recurrent if it overlapped ≥ 40% with one of the 85 CNVs and/or included the key genes of corresponding region (see details for each recurrent CNV in Supplementary Table 1).

### Deep phenotyping using behavioral, cognitive, and socio-demographic data

We analyzed a rich battery of cognitive, socio-demographic and environmental data partially reported in prior research^15^. Tabulated data of 1,319 phenotypes measured at baseline from 11,879 participants were imported and processed using Python. In line with previous research^15^, we used robust z-scores for the preprocessing of each phenotype. The robust z-scores are derived by calculating each phenotype’s absolute deviation from the median absolute deviation (MAD)^73^. In other words, the resulting score indicates how many standard deviations each value deviates from the median, with robustness to outliers. Subsequently, we removed values with a z-score > 4 (4 × MAD). We then excluded phenotypes with less than 75% retained values before excluding subjects with less than 75 % retained values across the retained phenotypes. The remaining subjects (n = 11,867) were considered for further analysis. Finally, all data included in the study were visually checked by the same researcher (JK). The complete list of 962 phenotypes from 20 predefined categories included in the analysis is available in Table S1. As the last step, for the purpose of data analysis, missing values were imputed using the KNNImputer function (n_neighbors = 5, weights = “uniform”) from the scikit-learn package. All derived phenotypic measures were then adjusted for variation that can be explained by age and sex.

### MRI imaging-derived phenotypes

Our data sample included expert-curated brain-imaging phenotypes of grey matter morphology (T1w MRI). The images were acquired across 21 sites in the United States with harmonized imaging protocols for GE, Philips, and Siemens scanners^74^. We used baseline structural T1-weighted tabulated MRI data from ABCD curated release 4.0. We only included participants who passed quality assurance using the recommended QC parameters (n = 11,723) described in the ABCD 4.0 Imaging Instruments Release Notes. ABCD preprocessing and QC steps are described in detail in the methodological reference for the ABCD study^74^.

Tabulated brain-imaging phenotypes were guided by the topographical brain region definitions based on the Destrieux parcellation atlas^75^. This feature-generation step provides neurobiologically interpretable measures of gray matter volume in 148 regions. For each included regional volume, we calculated the MAD for each brain region and removed values with MAD > 4^15^. Subjects with less than 90% of regional volume retained in any region were excluded from the analysis. The remaining subjects (n = 11,723) were included for further analysis. Finally, interindividual variations in the volumes that could be explained by nuisance variables outside primary interest were adjusted for by regressing out: age, sex, total brain volume, and scanning site.

We used the BrainStat toolbox^76^ to contextualize obtained patterns with respect to large-scale brain networks defined by Schaefer-Yeo definition^77^. Specifically, we mapped the brain loadings sizes from the 148 regions to fsaverage5 vertices, which we then averaged across the seven resting state brain networks.

To analyze temporal changes in brain structure, we also acquired structural T1-weighted tabulated MRI data during the follow-up two years after the first MRI recording. Brain imaging data from this second time point underwent the same cleaning steps as the baseline brain imaging data. In total, follow-up brain measurements were available for 5,663 subjects.

### Multivariate pattern analysis protocol

Our analysis aimed to reveal dominant modes of covariation between brain and behavior in the general population. Such a holistic perspective on complex brain-behavior relationships might be challenging to discern through univariate analyses. We leveraged 7,657 participants with both brain and behavior measurements that passed quality control. As a first data cleaning step, each brain and behavior measurement was normalized (z-scored) and submitted (separately for brain and behavior measurements) to principal component analysis (PCA) in order to increase the robustness of subsequent steps by avoiding potential issues with rank deficiency and fitting to noise. Based on a thorough examination, we extracted the first 100 PCA components for both the brain and behavior measurements (Sup. Fig. 6). It is noteworthy that although we used regional volumes as measures of brain structure, comparable results, especially for behavioral loadings, were obtained with regional thickness and area (Sup. Fig. 7).

After initial cleaning steps, we used measurements from the participants not carrying any CNV to reveal modes of covariation in the general population. Therefore, the first set of cleaned input variables comprised regional brain volumes (5,535 × 100 matrix) and the second input variable set was constructed from behavior measurement (5,535 × 100 matrix). We elected canonical partial least squares analysis (PLS) as a multivariate pattern learning approach that is ideally suited to delineate relationships between two high-dimensional sets of variables by identifying latent structures that maximize the covariance between them^78^. In other words, PLS involves finding canonical modes that maximize covariations between the weighted set (linear combination) of behavior measures and a weighted set (linear combination) of brain structure measures. These PLS modes are identified by solving the generalized eigenvalue problem of the brain-behavior cross-covariance matrix. The subject-wise latent variables (i.e., linear combinations of original measures) are called brain and behavior scores, respectively, throughout the manuscript. The ensuing set of PLS *k* orthogonal modes of variation were uncorrelated with each other and naturally ordered from the most to least important. Therefore, the first and strongest mode explained the largest fraction of covariance between brain and behavior measurements. Each ensuing mode captured a fraction of brain-behavior covariation not explained by one of the *k* − 1 other modes. Notably, PLS draws similarities with canonical correlation analysis (CCA). However, CCA can be prone to instability^79^. Nevertheless, our obtained PLS solutions strongly resemble those obtained with CCA (Sup. Fig. 8).

### Contribution of original phenotypes to latent variables

In order to quantify the contribution of each phenotype (e.g., regional volume or behavioral measure) to the construction of the latent structures, we computed PLS loading. These loadings are obtained as Pearson’s correlation between a respective PLS score and the original measurement across subjects. The loadings thus indicate the strength and direction of the relationship between the original phenotype and the identified canonical components. Stronger loading values signify greater importance in contributing to the latent structures, offering insights into which variables drive the covariation patterns between brain architecture and behavior.

As a test for significance, above-chance phenotype contribution, we embraced a bootstrapping resampling strategy for the PLS model. In the first phase, a randomly perturbed version of the dataset was created by sampling controls with the same sample size with replacement. We repeated the bootstrap resampling procedure with 1,000 iterations. In so doing, we obtained different realizations of the entire analysis workflow and ensuing PLS model estimates. Concretely, the bootstrapping algorithm resulted in 1,000 instances of the trained PLS models used to obtain 1,000 sets of associated PLS coefficients.

Statistically salient coefficients had a distribution of 1,000 PLS coefficients significantly different from 0. Specifically, they were robustly different from zero if their two-sided confidence interval according to the 2.5/97.5% bootstrap-derived distribution did not include zero coefficient value, indicating the absence of an effect.

### Optimal number of PLS dimensions in the general population

Each identified PLS mode was entered into statistical significance tests of robustness consistent with an established combination of cross-validation and permutation testing^15^ (Sup. Fig. 1). Initially, controls are split into 10 folds, where nine folds of participants are used as a train set, and one fold is used as a test set. The controls in the training set are used to estimate the parameters of all subsequent tools. In the first step, each brain or behavior measurement is z-scored column-wise across all controls in the training set. PCA then separately reduces the dimension of brain and behavior measurements to 100 features. In the next step, the behavior and brain measures are used as input variables to estimate a single multivariate canonical PLS model, where the output of the model is a set of scores (latent variables). This PLS model can also be characterized by weights (projection matrices used to transform input variables). PLS weights are back-projected using the PCA model to obtain brain and behavior weights in the original non-reduced ambient space.

In the next step, brain and behavior scores are computed for controls from the test set. Specifically, z-scoring followed by PCA dimensionality reduction is applied with parameters learned using the training set. The resulting preprocessed measurements are multiplied by PLS weights to obtain PLS scores for test-sample controls. Finally, the covariance between brain and behavior scores is calculated for each canonical mode. We took the average of these canonical covariances across the 10 folds. This procedure is repeated 100 times with a random fold split of controls to obtain a distribution of out-of-sample covariances for each PLS mode.

To assess the significance of the resulting PLS modes, we ran 1,000 iterations of the same 10-fold cross-validation procedure described above, where the order of participants of the brain measurements was randomly permuted in each iteration. In contrast to the unpermuted dataset, we collected covariances for the training rather than the testing subjects to account for overfitting by the PLS. In other words, using covariance from the permuted train set, and not the test set, represents a more stringent criterion. Finally, P values for each of the PLS modes were calculated as a percentage of cases when permuted covariance was greater than the mean cross-validated covariance.

### Group differences in the expression of brain and behavior patterns

One of the main objectives of our analyses was to investigate the effect of CNV carriership on the revealed modes of brain-behavior covariation. To that end, we developed a pipeline quantifying the differences in brain and behavior scores for the identified significant PLS mode (Sup. Fig. 1). Initially, participants without any CNV are split into a training set (80%) and a test set (20%). As described above, brain and behavior scores are computed for controls from the test set using parameters learned from the training set. In addition, brain and behavior scores are also calculated for CNV carriers (either deletion or duplication). Specifically, z-scoring followed by PCA dimensionality reduction is applied with parameters learned using the training set of controls. The resulting preprocessed measurements are multiplied by PLS weights to obtain PLS scores for CNV carriers. Finally, the differences in both brain and behavior scores between out-of-sample controls and CNV carriers are calculated for each canonical mode. This procedure is repeated 1,000 times with a random 80:20 split of controls to obtain a distribution of PLS score differences. Finally, P values for each of the PLS modes were calculated as a percentage of cases when the difference between mean scores of CNV carriers and mean scores of out-of-sample controls was greater than zero (resp. lower for modes with negative mean expression).

### Temporal shift in brain pattern expressions

In total, 3,654 participants passed the quality control of genetic data and had brain recordings measured at the baseline and 2-year follow-up. For these subjects, we compared brain scores from a single PLS model that subsequently evaluated brain baseline and follow-up measurements. In the first step, the parameters of a PLS model were estimated using baseline measurements of brain and behavior. Using these brain loadings, we then calculated a brain score for each participant in each visit across the significant PLS modes. Furthermore, we used a bootstrap mean test to evaluate if there was a significant temporal change in brain scores in controls and CNV carriers. In this non-parametric approach to statistical inference, 1,000 bootstrap samples are drawn with replacements from the brain scores. The mean is then calculated for each of these bootstrap samples, creating a distribution of sample means for both sets of brain scores. Finally, there is a significant difference in PLS scores between baseline and follow-up if the two-sided confidence interval according to the 2.5/97.5% distribution of 1,000 differences does not include zero.

### Effect size of CNV carriership

We used Cohen’s d to quantify the effect size of the CNVs on revealed PLS modes. For a given mode and separately for brain and behavior, Cohen’s d is calculated as:

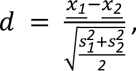

where <u>*x_1_*</u> corresponds to the mean PLS score across CNV carriers, <u>*x_2_*</u> corresponds to the mean PLS score across controls. Similarly, s1 and s2 correspond to standard deviations of PLS scores of CNV carriers and controls.

## Data availability

The data supporting the findings of this study are available from the Adolescent Brain Cognitive Development (ABCD) dataset. The ABCD dataset is a publicly available resource accessible through the National Institute of Mental Health Data Archive (NDA). The specific data used in this study can be located within the ABCD dataset under X.

## Code availability

The processing scripts and custom analysis software used in this work are available in a publicly accessible GitHub repository, along with examples of key visualizations in the paper: https://github.com/dblabs-mcgill-mila/CNV-covariation.

## Acknowledgements

DB was supported by the Brain Canada Foundation, through the Canada Brain Research Fund, with the financial support of Health Canada, National Institutes of Health (NIH R01 AG068563A, NIH R01 DA053301-01A1, NIH R01 MH129858-01A1), the Canadian Institute of Health Research (CIHR 438531, CIHR 470425), the Healthy Brains Healthy Lives initiative (Canada First Research Excellence fund), the IVADO R^3^AI initiative (Canada First Research Excellence fund), and by the CIFAR Artificial Intelligence Chairs program (Canada Institute for Advanced Research). This research was also supported by Calcul Quebec (http://www.calculquebec.ca) and Compute Canada (http://www.computecanada.ca), the Brain Canada Multi-Investigator initiative, the Canadian Institutes of Health Research, CIHR_400528, The Institute of Data Valorization (IVADO) through the Canada First Research Excellence Fund, Healthy Brains for Healthy Lives through the Canada First Research Excellence Fund. SJ is a recipient of a Canada Research Chair in neurodevelopmental disorders and a chair from the Jeanne et Jean Louis Levesque Foundation. KK was supported by The Institute of Data Valorization (IVADO) Postdoctoral Fellowship program through the Canada First Research Excellence Fund. BTTY is supposed by the NUS Yong Loo Lin School of Medicine (NUHSRO/2020/124/TMR/LOA), the Singapore National Medical Research Council (NMRC) LCG (OFLCG19May-0035), NMRC CTG-IIT (CTGIIT23jan-0001), NMRC STaR (STaR20nov-0003), Singapore Ministry of Health (MOH) Centre Grant (CG21APR1009), the Temasek Foundation (TF2223-IMH-01), and the United States National Institutes of Health (R01MH120080 & R01MH133334). The funders had no role in study design, data collection and analysis, the decision to publish or the preparation of the manuscript.

## Author Contributions Statement

DB and JK designed the study, analyzed brain and behavioral data, and drafted the manuscript. SJ, GH, ZS, and MJL called CNVs. DB and SJ contributed to the interpretation of the results and in the editing of the manuscript. All authors provided feedback on the manuscript. DB led data analytics.

## Competing Interests Statement

DB is a shareholder and advisory board member at MindState Design Labs, USA. OAA is a consultant to Cortechs.ai. PT obtained grant support from Biogen, Inc., for research unrelated to this manuscript.

## Notes

### Author Declarations

The data supporting the findings of this study are available from the Adolescent Brain Cognitive Development (ABCD) dataset. The ABCD dataset is a publicly available resource accessible through the National Institute of Mental Health Data Archive (NDA).

## References

1. Thompson, P. M. et al. Genetic influences on brain structure. Nat Neurosci 4, 1253–1258 (2001).

2. Timpson, N. J., Greenwood, C. M. T., Soranzo, N., Lawson, D. J. & Richards, J. B. Genetic architecture: the shape of the genetic contribution to human traits and disease. Nat Rev Genet 19, 110–124 (2018).

3. Claussnitzer, M. et al. A brief history of human disease genetics. Nature 577, 179–189 (2020).

4. Manolio, T. A. et al. Finding the missing heritability of complex diseases. Nature 461, 747–753 (2009).

5. Conrad, D. F. et al. Origins and functional impact of copy number variation in the human genome. Nature 464, 704–712 (2010).

6. Freeman, J. L. et al. Copy number variation: New insights in genome diversity. Genome Res. 16, 949–961 (2006).

7. Auwerx, C. et al. The individual and global impact of copy-number variants on complex human traits. Am J Hum Genet S0002–9297(22)00061–1 (2022) doi:10.1016/j.ajhg.2022.02.010.

8. Kopal, J. et al. Rare CNVs and phenome-wide profiling highlight brain structural divergence and phenotypical convergence. Nat Hum Behav 1–17 (2023) doi:10.1038/s41562-023-01541-9.

9. Chawner, S. J. R. A. et al. Genotype-phenotype associations in children with copy number variants associated with high neuropsychiatric risk in the UK (IMAGINE-ID): a case-control cohort study. Lancet Psychiatry 6, 493–505 (2019).

10. Marshall, C. R. et al. Contribution of copy number variants to schizophrenia from a genome-wide study of 41,321 subjects. Nat Genet 49, 27–35 (2017).

11. Sanders, S. J. et al. A framework for the investigation of rare genetic disorders in neuropsychiatry. Nat Med 25, 1477–1487 (2019).

12. Fuhrmann, D., Knoll, L. J. & Blakemore, S.-J. Adolescence as a Sensitive Period of Brain Development. Trends Cogn Sci 19, 558–566 (2015).

13. Tost, H., Champagne, F. A. & Meyer-Lindenberg, A. Environmental influence in the brain, human welfare and mental health. Nat Neurosci 18, 1421–1431 (2015).

14. Paus, T., Keshavan, M. & Giedd, J. N. Why do many psychiatric disorders emerge during adolescence? Nat Rev Neurosci 9, 947–957 (2008).

15. Alnæs, D., Kaufmann, T., Marquand, A. F., Smith, S. M. & Westlye, L. T. Patterns of sociocognitive stratification and perinatal risk in the child brain. Proceedings of the National Academy of Sciences 117, 12419–12427 (2020).

16. Dahl, R. E., Allen, N. B., Wilbrecht, L. & Suleiman, A. B. Importance of investing in adolescence from a developmental science perspective. Nature 554, 441–450 (2018).

17. Millan, M. J. et al. Altering the course of schizophrenia: progress and perspectives. Nat Rev Drug Discov 15, 485–515 (2016).

18. Casey, B. J. et al. The Adolescent Brain Cognitive Development (ABCD) study: Imaging acquisition across 21 sites. Developmental Cognitive Neuroscience 32, 43–54 (2018).

19. Smith, S. M. et al. A positive-negative mode of population covariation links brain connectivity, demographics and behavior. Nature Neuroscience 18, 1565–1567 (2015).

20. Stalnaker, T. A., Cooch, N. K. & Schoenbaum, G. What the orbitofrontal cortex does not do. Nat Neurosci 18, 620–627 (2015).

21. Modenato, C. et al. Effects of eight neuropsychiatric copy number variants on human brain structure. Transl Psychiatry 11, 1–10 (2021).

22. Hartwigsen, G. et al. Phonological decisions require both the left and right supramarginal gyri. Proc Natl Acad Sci U S A 107, 16494–16499 (2010).

23. Mechelli, A., Humphreys, G. W., Mayall, K., Olson, A. & Price, C. J. Differential effects of word length and visual contrast in the fusiform and lingual gyri during reading. Proc Biol Sci 267, 1909–1913 (2000).

24. Vossel, S., Geng, J. J. & Fink, G. R. Dorsal and Ventral Attention Systems. Neuroscientist 20, 150–159 (2014).

25. Buckner, R. L., Andrews-Hanna, J. R. & Schacter, D. L. The brain’s default network: anatomy, function, and relevance to disease. Ann N Y Acad Sci 1124, 1–38 (2008).

26. Kopal, J., Uddin, L. Q. & Bzdok, D. The end game: respecting major sources of population diversity. Nat Methods 20, 1122–1128 (2023).

27. Kopal, J. et al. Using rare genetic mutations to revisit structural brain asymmetry. bioRxiv 2023.04.17.537199 (2023) doi:10.1101/2023.04.17.537199.

28. Bzdok, D. Classical Statistics and Statistical Learning in Imaging Neuroscience. Front. Neurosci. 0, (2017).

29. Smith, S. M. & Nichols, T. E. Statistical Challenges in ‘Big Data’ Human Neuroimaging. Neuron 97, 263–268 (2018).

30. Bzdok, D., Nichols, T. E. & Smith, S. M. Towards Algorithmic Analytics for Large-scale Datasets. Nat Mach Intell 1, 296–306 (2019).

31. Bzdok, D. & Ioannidis, J. P. A. Exploration, Inference, and Prediction in Neuroscience and Biomedicine. Trends in Neurosciences 42, 251–262 (2019).

32. Nicolaisen-Sobesky, E. et al. A cross-cohort replicable and heritable latent dimension linking behaviour to multi-featured brain structure. Commun Biol 5, 1–11 (2022).

33. Cooper, G. M. et al. A Copy Number Variation Morbidity Map of Developmental Delay. Nat Genet 43, 838–846 (2011).

34. Huguet, G. et al. Measuring and Estimating the Effect Sizes of Copy Number Variants on General Intelligence in Community-Based Samples. JAMA Psychiatry 75, 447–457 (2018).

35. Kim, S. H. et al. Language characterization in 16p11.2 deletion and duplication syndromes. Am J Med Genet B Neuropsychiatr Genet 183, 380–391 (2020).

36. Kendall, K. M. et al. Cognitive performance and functional outcomes of carriers of pathogenic copy number variants: analysis of the UK Biobank. Br J Psychiatry 214, 297–304 (2019).

37. Mollon, J., Almasy, L., Jacquemont, S. & Glahn, D. C. The contribution of copy number variants to psychiatric symptoms and cognitive ability. Mol Psychiatry 1–14 (2023) doi:10.1038/s41380-023-01978-4.

38. Wang, H.-T. et al. Finding the needle in a high-dimensional haystack: Canonical correlation analysis for neuroscientists. Neuroimage 216, 116745 (2020).

39. McCarroll, S. A. et al. Integrated detection and population-genetic analysis of SNPs and copy number variation. Nat Genet 40, 1166–1174 (2008).

40. Morris, T. T., Davies, N. M., Hemani, G. & Smith, G. D. Population phenomena inflate genetic associations of complex social traits. Sci Adv 6, eaay0328 (2020).

41. Benkarim, O. et al. Population heterogeneity in clinical cohorts affects the predictive accuracy of brain imaging. PLOS Biology 20, e3001627 (2022).

42. Glasser, M. F. & Van Essen, D. C. Mapping Human Cortical Areas In Vivo Based on Myelin Content as Revealed by T1- and T2-Weighted MRI. J Neurosci 31, 11597–11616 (2011).

43. Gogtay, N. et al. Dynamic mapping of human cortical development during childhood through early adulthood. Proc Natl Acad Sci U S A 101, 8174–8179 (2004).

44. Schulz, A. & Northridge, M. E. Social determinants of health: implications for environmental health promotion. Health Educ Behav 31, 455–471 (2004).

45. Pickett, K. E. & Pearl, M. Multilevel analyses of neighbourhood socioeconomic context and health outcomes: a critical review. J Epidemiol Community Health 55, 111–122 (2001).

46. Robinette, J. W., Charles, S. T. & Gruenewald, T. L. Neighborhood Socioeconomic Status and Health: A Longitudinal Analysis. J Community Health 42, 865–871 (2017).

47. Robinette, J. W., Piazza, J. R. & Stawski, R. S. Neighborhood safety concerns and daily well-being: A national diary study. Wellbeing, space and society 2, (2021).

48. Martin-Brevet, S. et al. Quantifying the Effects of 16p11.2 Copy Number Variants on Brain Structure: A Multisite Genetic-First Study. Biological Psychiatry 84, 253–264 (2018).

49. Modenato, C. et al. Lessons Learned From Neuroimaging Studies of Copy Number Variants: A Systematic Review. Biological Psychiatry 90, 596–610 (2021).

50. Ulfarsson, M. O. et al. 15q11.2 CNV affects cognitive, structural and functional correlates of dyslexia and dyscalculia. Transl Psychiatry 7, e1109 (2017).

51. Couvy-Duchesne, B. et al. Lingual Gyrus Surface Area Is Associated with Anxiety-Depression Severity in Young Adults: A Genetic Clustering Approach. eNeuro 5, ENEURO.0153-17.2017 (2018).

52. Japee, S., Holiday, K., Satyshur, M. D., Mukai, I. & Ungerleider, L. G. A role of right middle frontal gyrus in reorienting of attention: a case study. Front Syst Neurosci 9, 23 (2015).

53. Alexander-Bloch, A. et al. Copy Number Variant Risk Scores Associated With Cognition, Psychopathology, and Brain Structure in Youths in the Philadelphia Neurodevelopmental Cohort. JAMA Psychiatry 79, 699–709 (2022).

54. Volkow, N. D. et al. The conception of the ABCD study: From substance use to a broad NIH collaboration. Dev Cogn Neurosci 32, 4–7 (2018).

55. Auchter, A. M. et al. A description of the ABCD organizational structure and communication framework. Dev Cogn Neurosci 32, 8–15 (2018).

56. Paul, S. E. et al. Associations Between Prenatal Cannabis Exposure and Childhood Outcomes: Results From the ABCD Study. JAMA Psychiatry 78, 64–76 (2021).

57. Garavan, H. et al. Recruiting the ABCD sample: Design considerations and procedures. Dev Cogn Neurosci 32, 16–22 (2018).

58. Fan, C. C., Loughnan, R., Wilson, S., Hewitt, J. K., & ABCD Genetic Working Group. Genotype Data and Derived Genetic Instruments of Adolescent Brain Cognitive Development Study® for Better Understanding of Human Brain Development. Behav Genet 53, 159–168 (2023).

59. Chang, C. C. et al. Second-generation PLINK: rising to the challenge of larger and richer datasets. Gigascience 4, 7 (2015).

60. Huguet, G. et al. Genome-wide analysis of gene dosage in 24,092 individuals estimates that 10,000 genes modulate cognitive ability. Mol Psychiatry 26, 2663–2676 (2021).

61. Wang, K. et al. PennCNV: an integrated hidden Markov model designed for high-resolution copy number variation detection in whole-genome SNP genotyping data. Genome Res 17, 1665–1674 (2007).

62. Colella, S. et al. QuantiSNP: an Objective Bayes Hidden-Markov Model to detect and accurately map copy number variation using SNP genotyping data. Nucleic Acids Res 35, 2013–2025 (2007).

63. Purcell, S. et al. PLINK: A Tool Set for Whole-Genome Association and Population-Based Linkage Analyses. Am J Hum Genet 81, 559–575 (2007).

64. Karczewski, K. J. et al. The mutational constraint spectrum quantified from variation in 141,456 humans. Nature 581, 434–443 (2020).

65. Werling, D. M. et al. Whole-Genome and RNA Sequencing Reveal Variation and Transcriptomic Coordination in the Developing Human Prefrontal Cortex. Cell Reports 31, 107489 (2020).

66. Kang, H. J. et al. Spatio-temporal transcriptome of the human brain. Nature 478, 483–489 (2011).

67. Satterstrom, F. K. et al. Large-Scale Exome Sequencing Study Implicates Both Developmental and Functional Changes in the Neurobiology of Autism. Cell 180, 568–584.e23 (2020).

68. Schizophrenia Working Group of the Psychiatric Genomics Consortium. Biological insights from 108 schizophrenia-associated genetic loci. Nature 511, 421–427 (2014).

69. Coe, B. P. et al. Refining analyses of copy number variation identifies specific genes associated with developmental delay. Nat Genet 46, 1063–1071 (2014).

70. Stefansson, H. et al. CNVs conferring risk of autism or schizophrenia affect cognition in controls. Nature 505, 361–366 (2014).

71. Moreno-De-Luca, D. et al. Using large clinical data sets to infer pathogenicity for rare copy number variants in autism cohorts. Mol Psychiatry 18, 1090–1095 (2013).

72. Welcome to ClinGen. https://clinicalgenome.org/.

73. Leys, C., Ley, C., Klein, O., Bernard, P. & Licata, L. Detecting outliers: Do not use standard deviation around the mean, use absolute deviation around the median. Journal of Experimental Social Psychology 49, 764–766 (2013).

74. Hagler, D. J. et al. Image processing and analysis methods for the Adolescent Brain Cognitive Development Study. Neuroimage 202, 116091 (2019).

75. Destrieux, C., Fischl, B., Dale, A. & Halgren, E. Automatic parcellation of human cortical gyri and sulci using standard anatomical nomenclature. Neuroimage 53, 1–15 (2010).

76. Larivière, S. et al. BrainStat: A toolbox for brain-wide statistics and multimodal feature associations. NeuroImage 266, 119807 (2023).

77. Schaefer, A. et al. Local-Global Parcellation of the Human Cerebral Cortex from Intrinsic Functional Connectivity MRI. Cereb Cortex 28, 3095–3114 (2018).

78. Wegelin, J. A Survey of Partial Least Squares (PLS) Methods, with Emphasis on the Two-Block Case. Technical report (2000).

79. Mihalik, A. et al. Canonical Correlation Analysis and Partial Least Squares for Identifying Brain–Behavior Associations: A Tutorial and a Comparative Study. Biological Psychiatry: Cognitive Neuroscience and Neuroimaging 7, 1055–1067 (2022).

