## Supplementary material for "High-effect gene-coding variants impact cognition, mental well-being, and neighborhood safety substrates in brain morphology": Sup. Fig.

### Supplementary Figures

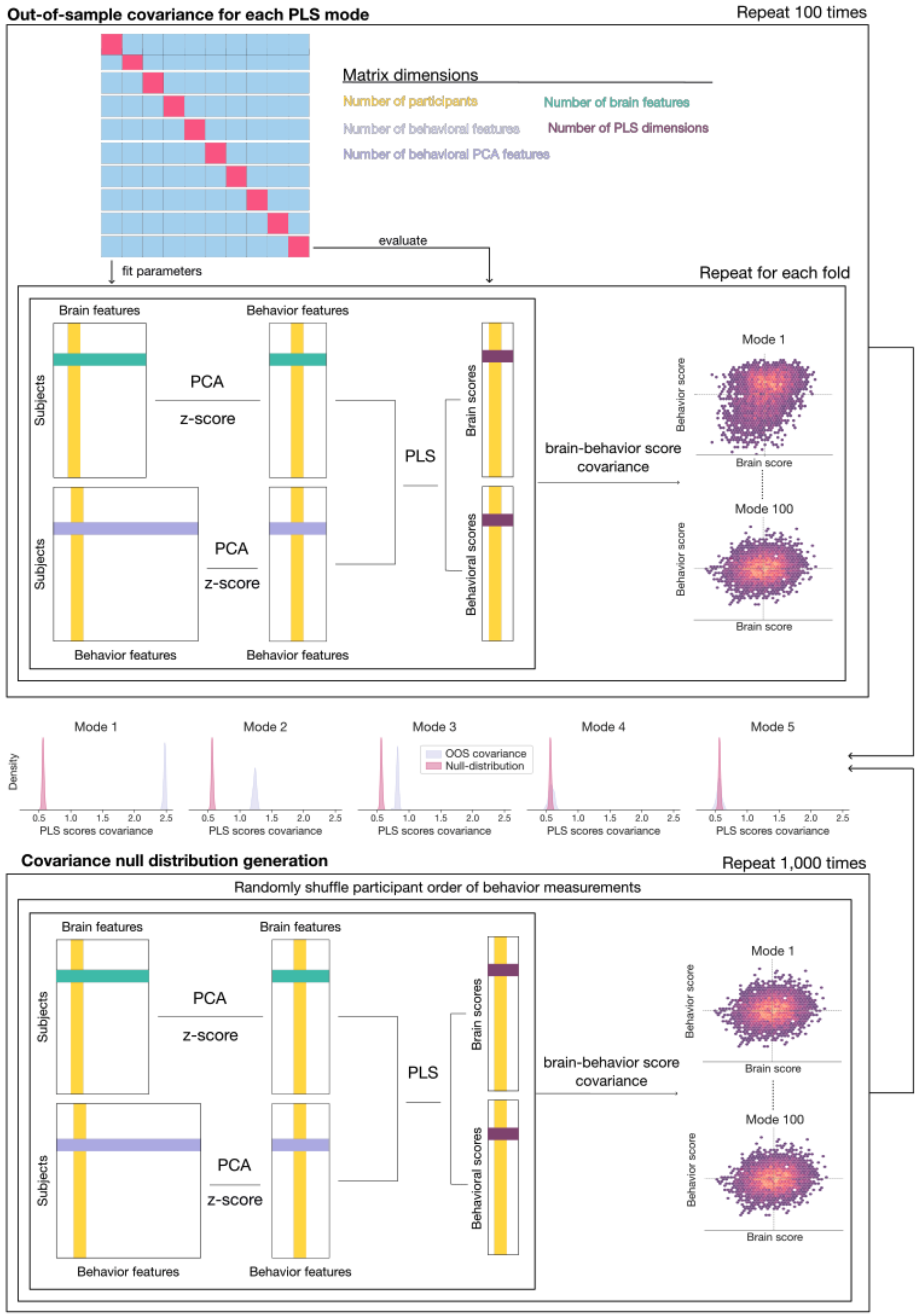


Supplementary Figure 1

**Schematic of selecting the optimal number of PLS dimensions in the general population**

The schematic depicts significance tests of revealed PLS using an established combination of cross-validation and permutation testing. The upper part shows the calculation of the distribution of out-of-sample covariances for each PLS mode. Initially, controls are split into 10 folds, where nine folds of participants are used as a train set, and one fold is used as a test set. The controls in the training set are used to estimate the parameters of all subsequent tools. In the first step, each brain or behavior measurement is z-scored column-wise across all controls in the training set. PCA then separately reduces the dimension of brain and behavior measurements to 100 features. In the next step, the behavior and brain measures are used as input variables to estimate a single multivariate canonical PLS model. PLS weights are back-projected using the PCA model to obtain brain and behavior weights in the original non-reduced ambient space. Afterwards, brain and behavior scores are computed for controls from the test set. Specifically, z-scoring followed by PCA dimensionality reduction is applied with parameters learned using the training set. The resulting preprocessed measurements are multiplied by PLS weights to obtain PLS scores for test-sample controls. Finally, the covariance between brain and behavior scores is calculated for each canonical mode. We took the average of these canonical covariances across the 10 folds. This procedure is repeated 100 times with a random fold split of controls. The bottom part shows the generation of covariance null distribution for each PLS mode. We ran 1,000 iterations of the same 10-fold cross-validation procedure described above, where the order of participants of the brain measurements was randomly permuted in each iteration. In contrast to the unpermuted dataset, we collected covariances for the training rather than the testing subjects to account for overfitting by the PLS. Finally, acknowledging familywise error, corrected P values for each of the PLS modes were calculated as a percentage of cases when permuted covariance was greater than the mean cross-validated covariance, as shown in the middle of the figure.

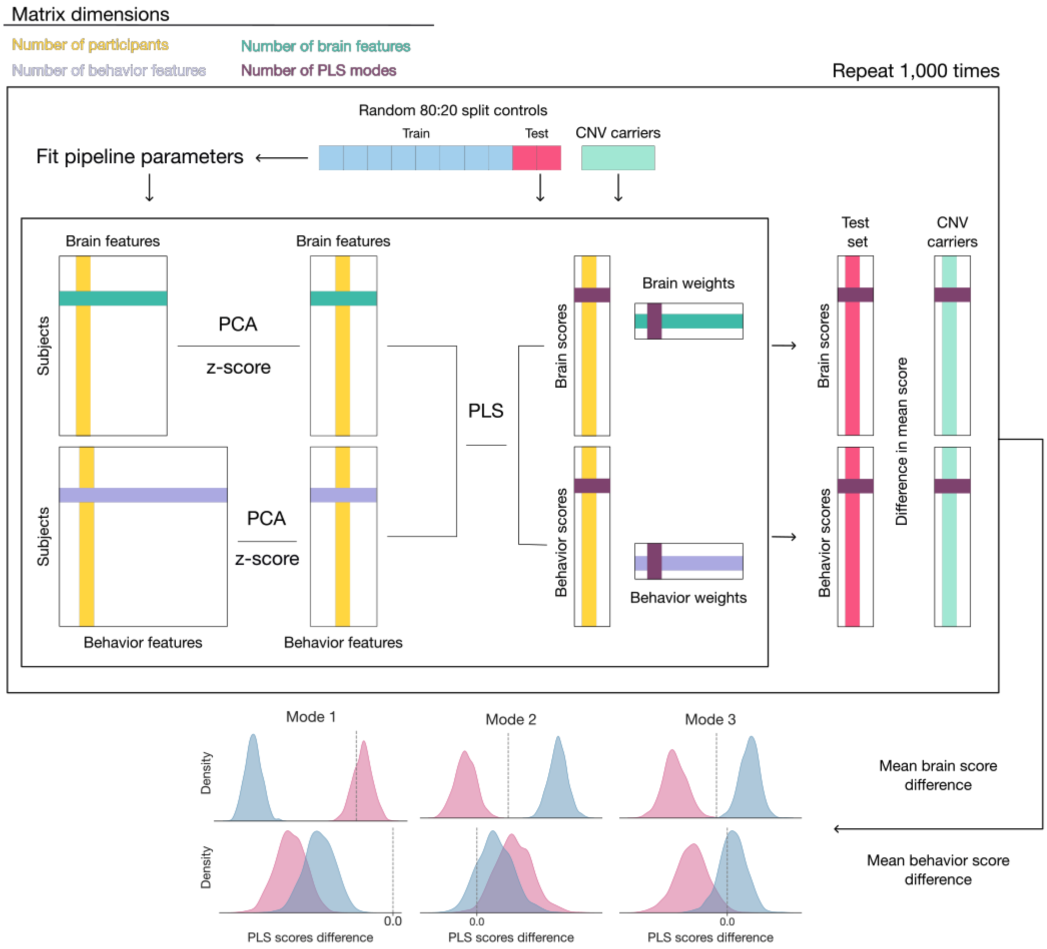


Supplementary Figure 2

**Schematic of quantifying the difference in PLS scores**

This figure illustrates the proposed pipeline evaluating the differences in brain and behavior scores for the three significant modes of population covariation between CNV carriers and out-of-sample controls. The upper part depicts the calculation of PLS scores for CNV carriers and test-sample controls. Initially, controls are split into a training set (80%) and a test set (20%). The controls in the training set are used to estimate the parameters of all subsequent tools. In the first step, each brain or behavior measurement is z-scored column-wise across all controls in the training set. PCA then reduces the dimension of behavior measurements to 300 features. In the next step, a single multivariate canonical PLS model is fitted to link measures of behavior and brain architecture. This PLS model can be characterized by a set of scores (latent variables) and weights (projection matrices used to transform original data). Behavior weights of the PLS model are back-projected using the PCA model to obtain behavior weights in the original non-reduced space. Now, brain and behavior scores are computed for CNV carriers and controls from the test set. Specifically, z-scoring followed by PCA dimensionality reduction is applied with parameters learned using the training set. The resulting preprocessed measurements are multiplied by PLS weights to obtain PLS scores for CNV carriers and test-sample controls. The bottom part illustrates the differences in both brain and behavior scores calculated for each canonical mode. This procedure is repeated 1,000 times with a random 80:20 split of controls to obtain a distribution of PLS score differences. There is a significant difference in PLS scores between CNV carriers and controls if the two-sided confidence interval according to the 2.5/97.5% distribution of 1,000 differences does not include zero.

Supplementary Figure 3

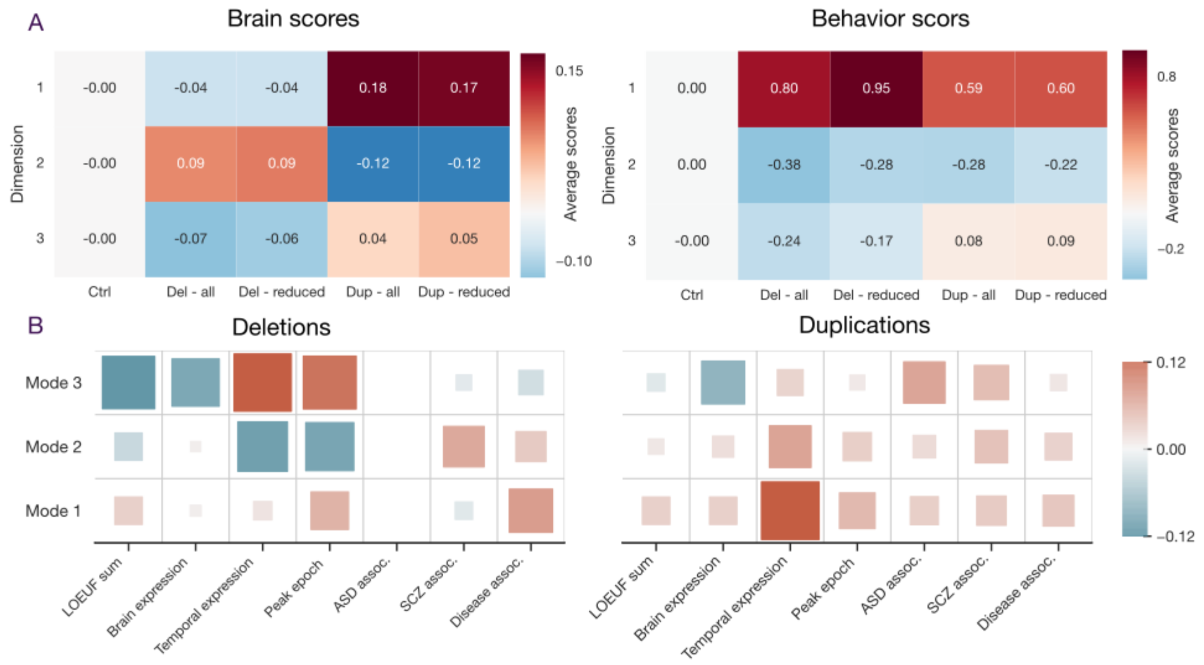


**Recurrent CNVs**

We conducted a sensitivity analysis to probe the effect of few recurrent CNVs (e.g., 1q21.2 or 16p11.2) on the observed differences in brain and behavior scores. To this end, we removed 100 carriers of recurrent deletions and 118 carriers of recurrent duplications. A) Effects on brain and behavior scores. The removal of participants carrying a recurrent CNV did not lead to quantitatively different behavioral brain scores across the three dimensions of population covariation. B) Annotation of genetic scores. We repeated an identical test the annotation of genes inside each CNV. We observe identical associations of CNV properties with behavioral scores as in the original sample Full list of recurrent CNVs in available as Supplementary Table 1.

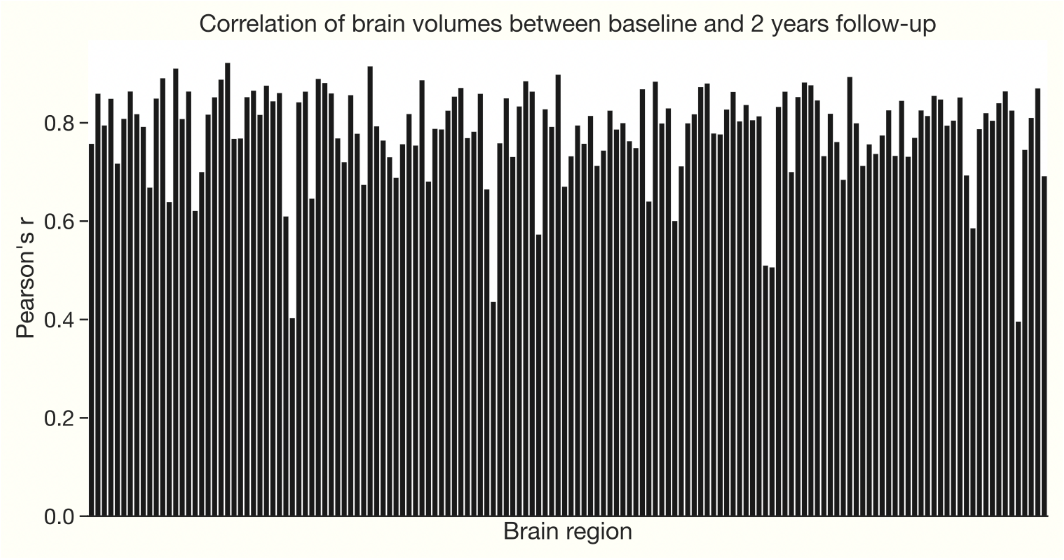


Supplementary Figure 4

**Similarity between baseline and 2-year follow-up brain measurements**

We quantified the similarity between regional brain volumes recorded at baseline and during follow-up measurement using Pearson's correlation. The bar plot depicts the linear association strength (y-axis) for each brain region (x-axis) across the 5,663 subjects.

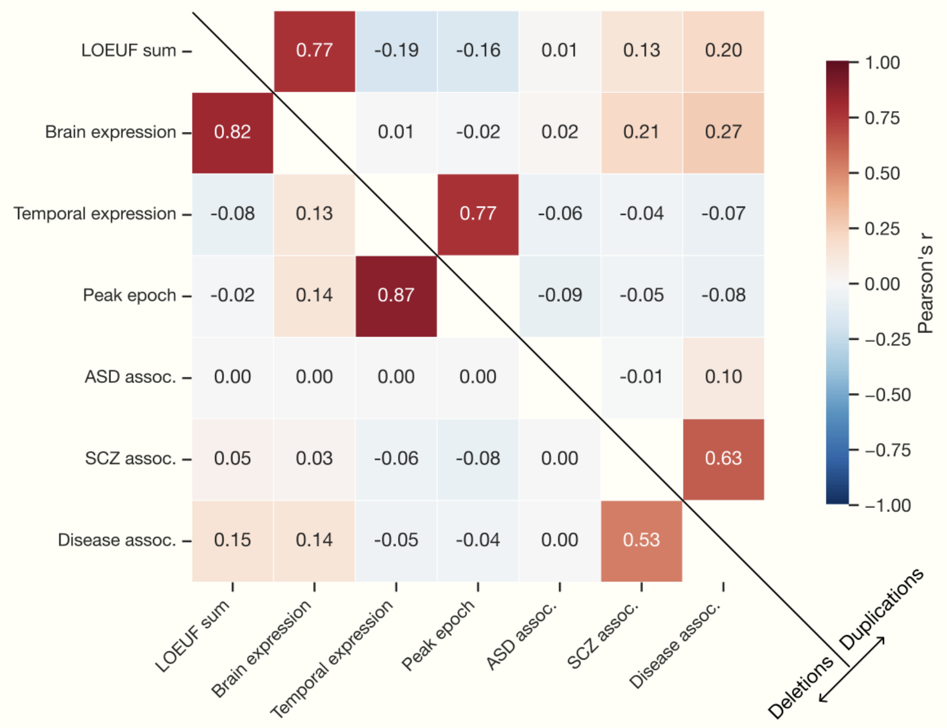


Supplementary Figure 5

**Correlation between genetic descriptors**

Each CNV carrier was annotated using seven descriptors characterizing the genetic content of the respective CNV. We here plot the correlation between these seven descriptors across deletion and duplication carriers. Especially the sum of LOEUF and the number of genes highly expressed in the brain (Pearson's r > 0.77) as well as temporal expression and peak epoch display very strong similarity (Pearson's r > 0.77).

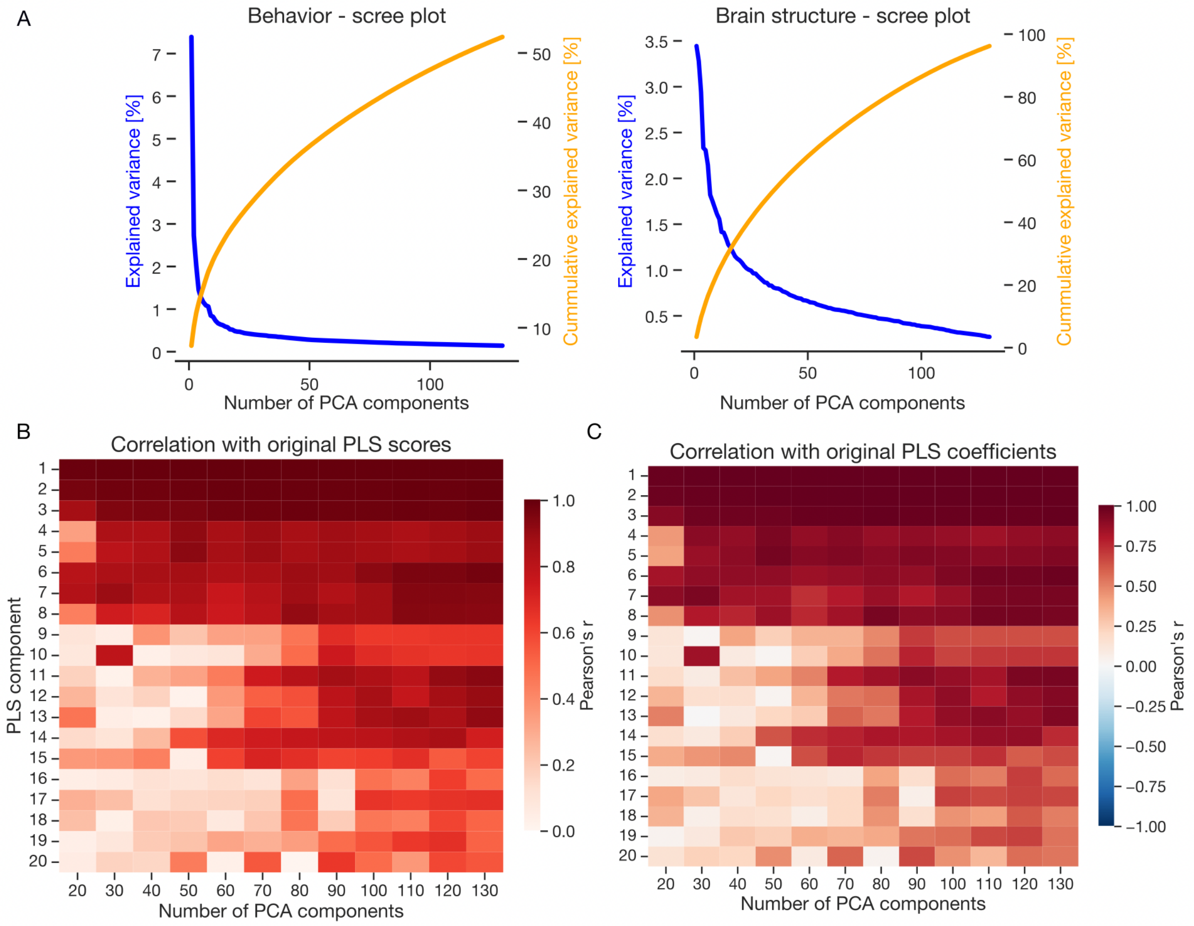


Supplementary Figure 6

**Number of optimal PCA components**

In order to avoid issues with rank deficiency and overfitting to noise, we reduced the dimension of original brain and behavior measurements using principal component analysis (PCA). A) Explained variance, as well as cumulative explained variance, is plotted separately for both brain and behavior measurements. Using the first 100 PCA components results in explaining 50 % of the behavioral variance and more than 95% variance in brain measurements. B) Comparison of PLS scores between PLS model with non-reduced and PCA-reduced inputs. We iteratively reduced the dimension of brain and behavior measurements using PCA. We then concatenated the resulting brain and behavior scores. Finally, concatenated scores were compared with the PLS score estimated using non-reduced measurements using Pearson's correlation. Using 100 PCA components results in stable results for the first 10 PLS dimensions. C) Comparison of PLS coefficients between PLS model with non-reduced and PCA-reduced inputs. This analysis is similar to the analysis of PLS scores with the exception that PLS coefficients derived after using PCA were back-projected in order to be comparable with original PLS coefficients. Obtained results strongly resembled results obtained from the analysis of PLS scores. Collectively, using 100 PCA components provides stable results while explaining a large partition of variance in the original measurements.

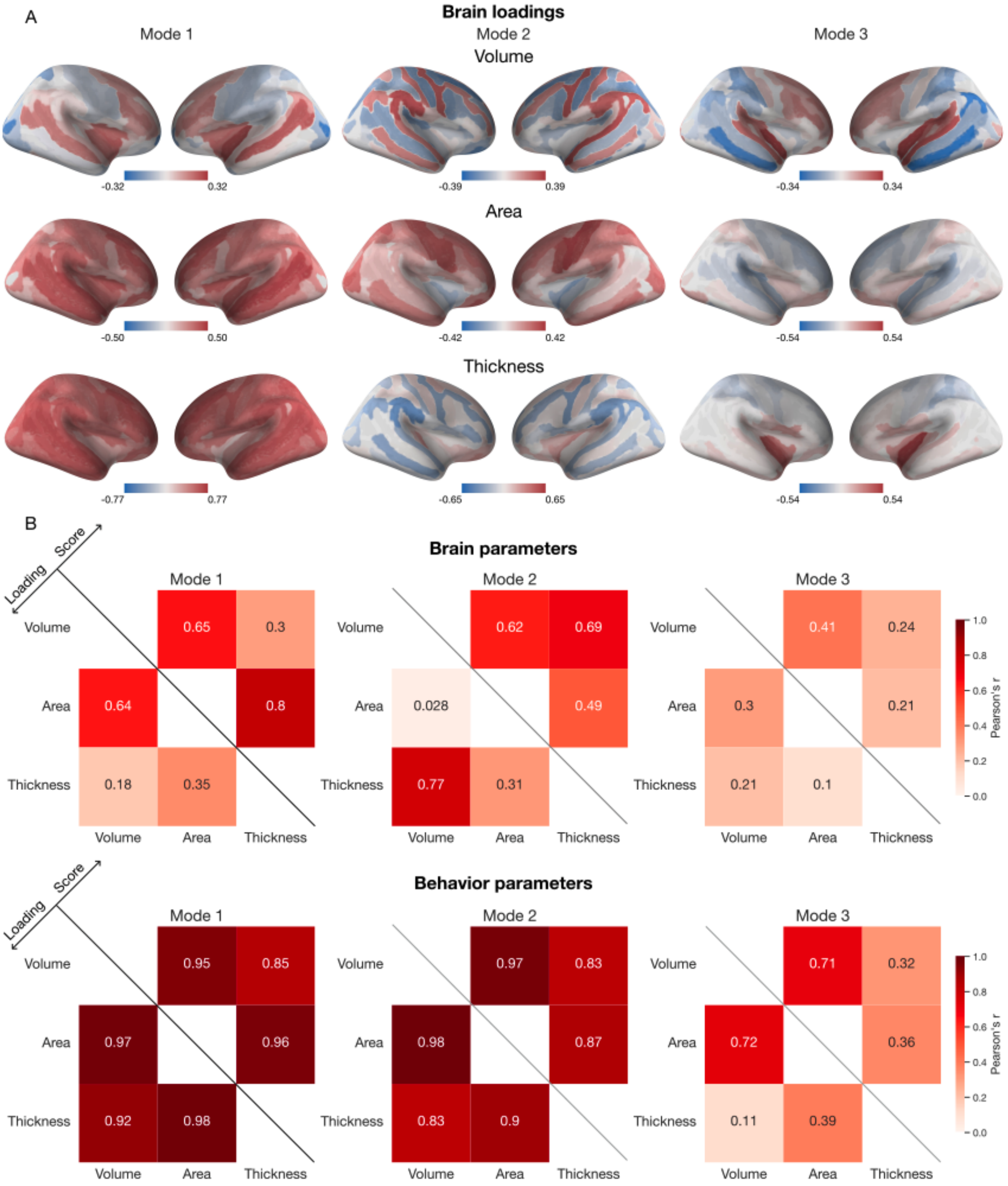


Supplementary Figure 7

**Comparison of PLS solutions based on regional volume, area, and thickness as a measure of brain architecture**

In the presented analyses, we uncovered hidden dimensions linking behavioral differentiation with brain architecture represented by regional brain volumes. We focused on a single measure of brain architecture in order to provide more interpretable results. Changes in brain volume can be indicative of structural alterations, such as atrophy or enlargement, which may be associated with various neurological or psychiatric conditions. To complement our analyses, we here provide supplementary analyses comparing our results with those obtained using regional area and thickness as measures of brain architecture. A) Brain loadings projected on the brain. Due to sign invariance, identical PLS models could carry opposite signs. B) In order to quantify the similarity between revealed brain patterns, we correlated regional brain loadings obtained using regional volume, area and thickness. In addition, we also compared behavioral loadings as well as brain and behavioral scores. The results of these analyses are summarized in the heatmaps. We observe strong similarity of PLS parameters among measures of brain architecture. Especially, behavior loadings and scores lead to almost identical solutions (Pearson's $r > 0.83$) for the first two PLS dimensions. Regarding brain parameters, loadings and scores based on volume most resemble those obtained using area in the first PLS dimension and thickness in the second PLS dimension.

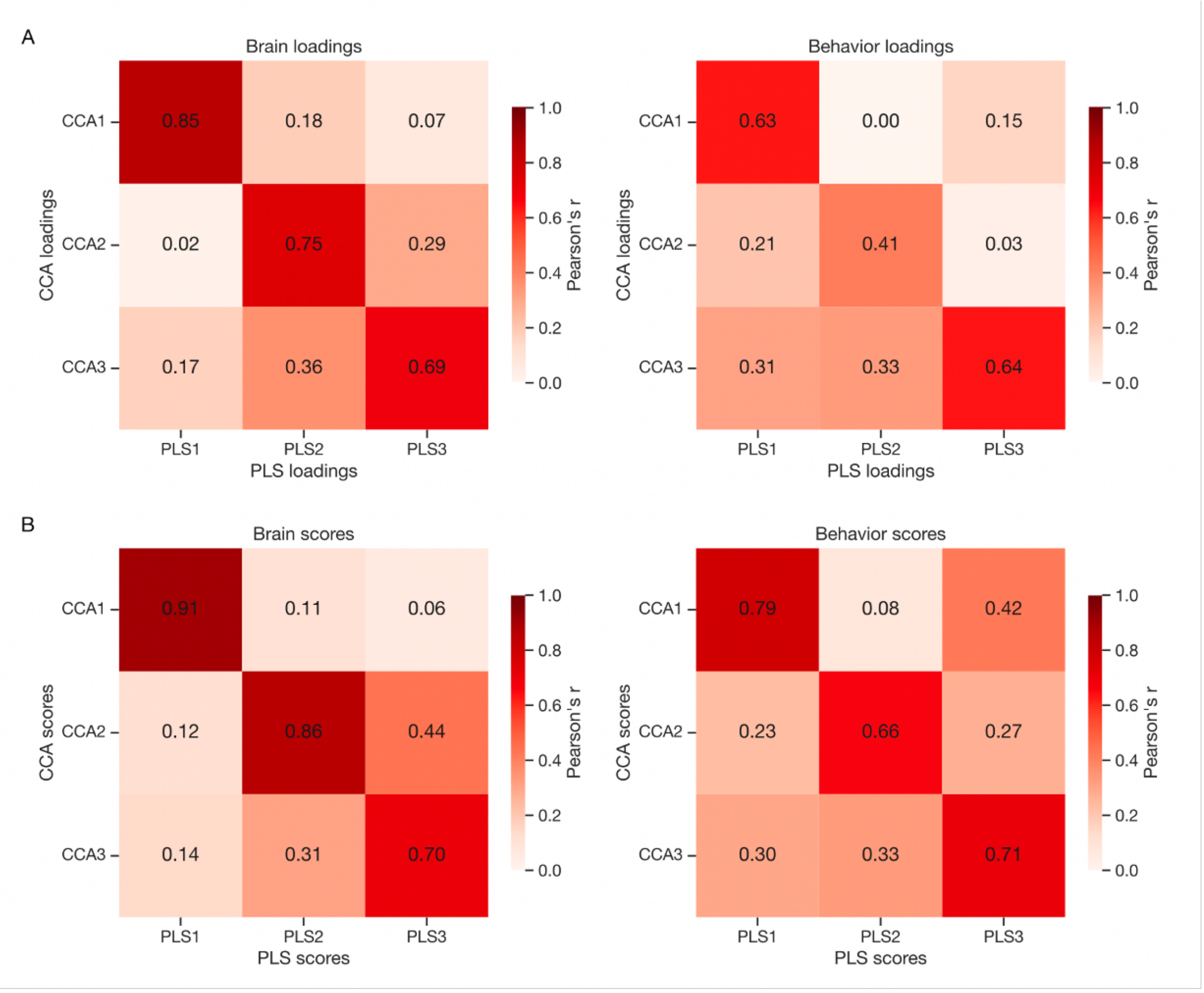


Supplementary Figure 8

**Canonical correlation analysis leads to quantitatively similar brain and behavior dimensions**

Canonical correlation analysis (CCA) and canonical partial least squares (PLS) are powerful multivariate methods for capturing associations across 2 modalities of data that share many attributes. In fact, CCA is a special case of PLS. CCA maximizes the correlation between the latent variables. CCA is thus more sensitive to the direction of the relationships across modalities and is not driven by within-modality variances. However, the optimization step in CCA is ill-posed (i.e., there is no unique solution) when the number of variables in at least one of the modalities exceeds the sample size. In addition, the CCA weights are unstable when the variables within one or both modalities are highly correlated. On the other hand, PLS maximizes covariance between latent variables. PLS is less sensitive to the direction of the across-modality relationships, as it is also driven by within-modality variances. Finally, PLS optimization is never ill-posed and copes with multicollinearity (i.e., standard PLS weights are stable)[75]. We compared our PLS solutions with those obtained using CCA. The heatmaps depict strong similarity between both loadings (A) and scores (B) for both brain and behavior variables between CCA and PLS.

### Supplementarry Tables

Supplementary Table 1 : List of recurrent CNV selected in the study based on Huguet et al 2021

| Chr. | Start | Stop | CNV | regionID | Clingen | Sum (1/LOEUF) | Condition to define recurrent CNV | | | |
|---|---|---|---|---|---|---|---|---|---|---|
| | | | | | | | Overlap | Size | Gene name | Gene note |
| chr1 | 1 | 2500000 | DEL | 1p36 | - | 75.6 | 40% | | *GABRD* | disrupted |
| chr1 | 145394955 | 145807817 | DEL | 1q21.1 TAR | - | 20.4 | 40% | <1 Mb | - | |
| chr1 | 145394955 | 145807817 | DUP | 1q21.1 TAR | - | 20.4 | 40% | <1 Mb | - | |
| chr1 | 145394955 | 147394444 | DEL | 1q21.1 distal+TAR | - | 30.6 | 40% | >=1 Mb | - | |
| chr1 | 145394955 | 147394444 | DUP | 1q21.1 distal + TAR | - | 30.6 | 40% | >=1 Mb | - | |
| chr1 | 146527987 | 147394444 | DEL | 1q21.1 distal | - | 9.1 | 40% | <1 Mb | - | |
| chr1 | 146527987 | 147394444 | DUP | 1q21.1 distal | - | 9.1 | 40% | <1 Mb | - | |
| chr2 | 50145643 | 51259674 | DEL | - | *NRXN1* | 3.9 | - | | *NRXN1* | disrupted-DEL /complete–DUP |
| chr2 | 96742409 | 97677516 | DEL | 2q11.2 | - | 43.0 | 40% | | *ARID5A; LMAN2L* | Disrupted |
| chr2 | 96742409 | 97677516 | DUP | 2q11.2 | - | 43.0 | 40% | | *ARID5A; LMAN2L* | disrupted |
| chr2 | 239716679 | 243199373 | DEL | 2q37 | *KIF1A* | 76.1 | 40% | | *HDAC4* | disrupted |
| chr2 | 239716679 | 243199373 | DUP | 2q37 | *KIF1A* | 76.1 | 40% | | *HDAC4* | disrupted |
| chr3 | 191517306 | 193017306 | DEL | 3q29 proximal | *FGF12* | 4.7 | 40% | | *FGF12* | disrupted |
| chr3 | 195720167 | 197354826 | DEL | 3q29 distal | - | 41.3 | 40% | | *DLG1* | disrupted |
| chr3 | 195720167 | 197354826 | DUP | 3q29 distal | - | 41.3 | 40% | | *DLG1* | disrupted |
| chr4 | 1552030 | 2091303 | DEL | 4p16.3(WH) | *NSD2* | 21.2 | 40% | | - | |
| chr4 | 1552030 | 2091303 | DUP | 4p16.3(WH) | *NSD2* | 21.2 | 40% | | - | |
| chr5 | 175720924 | 177052594 | DEL | 5q35 | *NSD1* | 68.0 | 40% | | - | |
| chr5 | 175720924 | 177052594 | DUP | 5q35 | *NSD1* | 68.0 | 40% | | - | |
| chr7 | 72744915 | 74142892 | DEL | 7q11.23 (WBS) | - | 50.2 | 40% | | *GTF2I; GTF2IRD 1* | disrupted |
| chr7 | 72744915 | 74142892 | DUP | 7q11.23 (WBS) | - | 50.2 | 40% | | *GTF2I; GTF2IRD 1* | disrupted |
| chr7 | 73978801 | 74144177 | DUP | 7q11.23 proximal | - | 6.7 | 40% | | - | |
| chr7 | 75138294 | 76064412 | DEL | 7q11.23 distal | - | 17.0 | 40% | | - | |
| chr8 | 8098990 | 11872558 | DEL | 8p23.1 | - | 25.2 | 40% | >=2 Mb | - | |
| chr8 | 8098990 | 11872558 | DUP | 8p23.1 | - | 25.2 | 40% | >=2 Mb | - | |
| chr9 | 140513444 | 140730578 | DEL | 9q34 *EHMT1* | *EHMT1* | 13.9 | 40% | >=1 Mb | *EHMT1* | disrupted |

| | | | | | | | | | | |
|---|---|---|---|---|---|---|---|---|---|---|
| chr9 | 140513444 | 140730578 | DUP | 9q34 *EHMT1* | *EHMT1* | 13.9 | 40% | >=1 Mb | *EHMT1* | disrupted |
| chr10 | 49390199 | 51058796 | DEL | 10q11.21q11.23 | - | 21.8 | 40% | | - | |
| chr10 | 49390199 | 51058796 | DUP | 10q11.21q11.23 | - | 21.8 | 40% | | - | |
| chr10 | 82045472 | 88931651 | DEL | 10q22q23 *NRG3; GRID1* | - | 41.1 | 40% | >=1 Mb | *GRID1; NRG3* | disrupted |
| chr10 | 82045472 | 88931651 | DUP | 10q22q23 *NRG3; GRID1* | - | 41.1 | 40% | >=1 Mb | *GRID1; NRG3* | disrupted |
| chr11 | 43940000 | 46020000 | DEL | 11p11.2 | - | 41.9 | 40% | | *EXT2* | disrupted |
| chr11 | 43940000 | 46020000 | DUP | 11p11.2 | - | 41.9 | 40% | | *EXT2* | disrupted |
| chr13 | 23555358 | 24884622 | DEL | 13q12.12 | - | 8.8 | 40% | | - | |
| chr13 | 23555358 | 24884622 | DUP | 13q12.12 | - | 8.8 | 40% | | - | |
| chr15 | 22805313 | 23094530 | DEL | 15q11.2 | - | 6.9 | 40% | | - | |
| chr15 | 22805313 | 23094530 | DUP | 15q11.2 | - | 6.9 | 40% | | - | |
| chr15 | 22805313 | 28390339 | DEL | 15q11.2q13.1 BP2-BP3 (PWS/AS) | *MAGEL2* | 39.8 | 40% | >=4 Mb | - | |
| chr15 | 22805313 | 28390339 | DUP | 15q11.2q13.1 BP2-BP3 (PWS/AS) | *MAGEL2* | 39.8 | 40% | >=4 Mb | - | |
| chr15 | 26971834 | 27548820 | DEL | 15q12 *GABRA5; GABRB3; GABRG3* | - | 8.2 | 40% | | *GABRA5; GABRB3; GABRG3* | disrupted |
| chr15 | 26971834 | 27548820 | DUP | 15q12 *GABRA5; GABRB3; GABRG3* | - | 8.2 | 40% | | *GABRA5 GABRB3 GABRG3* | disrupted |
| chr15 | 29161368 | 30375967 | DEL | 15q13.1q13.2 BP3-BP4 | - | 19.6 | 40% | | - | |
| chr15 | 29161368 | 30375967 | DUP | 15q13.1q13.2 BP3-BP4 | - | 19.6 | 40% | | - | |
| chr15 | 29161368 | 32462776 | DEL | 15q13.1q13.3 BP3-BP5 | - | 31.3 | 40% | | - | |
| chr15 | 29161368 | 32462776 | DUP | 15q13.1q13.3 BP3-BP5 | - | 31.3 | 40% | | - | |
| chr15 | 31080645 | 32462776 | DEL | 15q13.3 BP4-BP5 *CHRNA7* | - | 8.7 | 40% | | - | |
| chr15 | 31080645 | 32462776 | DUP | 15q13.3 BP4-BP5 *CHRNA7* | - | 8.7 | 40% | | - | |
| chr15 | 72900171 | 78151253 | DEL | 15q24 | - | 111.8 | 40% | >=1 Mb | - | |
| chr15 | 72900171 | 78151253 | DUP | 15q24 | - | 111.8 | 40% | >=1 Mb | - | |
| chr15 | 83219735 | 85722039 | DEL | 15q25.2 | - | 45.4 | 40% | >=1 Mb | - | |
| chr15 | 83219735 | 85722039 | DUP | 15q25.2 | - | 45.4 | 40% | >=1 Mb | - | |
| chr16 | 3775056 | 3930121 | DEL | - | *CREBBP* | 15.2 | - | | *CREBBP* | disrupted-DEL / completr-DUP |
| chr16 | 15511655 | 16293689 | DEL | 16p13.11 | - | 18.4 | 40% | | - | |

| | | | | | | | | | | |
|---|---|---|---|---|---|---|---|---|---|---|
| chr16 | 15511655 | 16293689 | DUP | 16p13.11 | - | 18.4 | 40% | | - | |
| chr16 | 21596415 | 28347808 | DEL | 16p11.2p12.1 | - | 112.3 | 40% | | - | |
| chr16 | 21596415 | 28347808 | DUP | 16p11.2p12.1 | - | 112.3 | 40% | | - | |
| chr16 | 21950135 | 22431889 | DEL | 16p12.1 | - | 9.0 | 40% | | - | |
| chr16 | 21950135 | 22431889 | DUP | 16p12.1 | - | 9.0 | 40% | | - | |
| chr16 | 28823196 | 29046783 | DEL | 16p11.2 distal | - | 21.4 | 40% | <1 Mb | - | |
| chr16 | 28823196 | 29046783 | DUP | 16p11.2 distal | - | 21.4 | 40% | <1 Mb | - | |
| chr16 | 28823196 | 30200773 | DEL | 16p11.2 distal+proximal | - | 62.6 | 40% | >=1 Mb | - | |
| chr16 | 28823196 | 30200773 | DUP | 16p11.2 distal+proximal | - | 62.6 | 40% | >=1 Mb | - | |
| chr16 | 29650840 | 30200773 | DEL | 16p11.2 proximal | - | 39.2 | 40% | <1 Mb | - | |
| chr16 | 29650840 | 30200773 | DUP | 16p11.2 proximal | - | 39.2 | 40% | <1 Mb | - | |
| chr17 | 2496923 | 2588909 | DEL | - | - | 9.3 | - | | *PAFAH1B1* | disrupted-DEL / complete-DUP |
| chr17 | 2496923 | 2588909 | DUP | - | - | 9.3 | - | | *PAFAH1B1* | disrupted-DEL / complete-DUP |
| chr17 | 14141387 | 15426961 | DEL | 17p12 (HNPP/CMT1A) | - | 6.5 | 40% | | *PMP22* | disrupted |
| chr17 | 14141387 | 15426961 | DUP | 17p12 (HNPP/CMT1A) | - | 6.5 | 40% | | *PMP22* | disrupted |
| chr17 | 16812771 | 20211017 | DEL | 17p11.2 (Potocki-Lupski/SMS) | *RAI1* | 74.1 | 40% | | - | |
| chr17 | 16812771 | 20211017 | DUP | 17p11.2 (Potocki-Lupski/SMS) | *RAI1* | 74.1 | 40% | | - | |
| chr17 | 29107491 | 30265075 | DEL | 17q11.2 *NF1* | - | 33.3 | 40% | | *NF1* | disrupted |
| chr17 | 29107491 | 30265075 | DUP | 17q11.2 *NF1* | - | 33.3 | 40% | | *NF1* | disrupted |
| chr17 | 34815904 | 36217432 | DEL | 17q12 *HNF1B* | - | 37.1 | 40% | | - | |
| chr17 | 34815904 | 36217432 | DUP | 17q12 *HNF1B* | - | 37.1 | 40% | | - | |
| chr17 | 43705356 | 44164691 | DEL | 17q21.31 | *KANSL1* | 8.3 | 40% | | - | |
| chr17 | 43705356 | 44164691 | DUP | 17q21.31 | *KANSL1* | 8.3 | 40% | | - | |
| chr17 | 58302389 | 60289141 | DEL | 17q23.1q23.2 | *PPM1D* | 56.8 | 40% | | - | |
| chr17 | 58302389 | 60289141 | DUP | 17q23.1q23.2 | *PPM1D* | 56.8 | 40% | | - | |
| chr22 | 19037332 | 21466726 | DEL | 22q11.2 proximal | - | 75.0 | 40% | | - | |
| chr22 | 19037332 | 21466726 | DUP | 22q11.2 proximal | - | 75.0 | 40% | | - | |
| chr22 | 21920127 | 23653646 | DEL | 22q11.2 distal | - | 30.0 | 40% | | - | |
| chr22 | 21920127 | 23653646 | DUP | 22q11.2 distal | - | 30.0 | 40% | | - | |
| chr22 | 51113070 | 51171640 | DEL | 22q13 *SHANK3* | *SHANK3* | 8.1 | 40% | >=1 Mb | *SHANK3* | disrupted |
| chr22 | 51113070 | 51171640 | DUP | 22q13 *SHANK3* | *SHANK3* | 8.1 | 40% | >=1 Mb | *SHANK3* | disrupted |
